# Development of a Core Outcome Set for Mild Cognitive Impairment (MCI-COS): Recommendations from a multistakeholder Delphi consensus study

**DOI:** 10.64898/2026.07.10.26357747

**Authors:** Victoria Grace Gabb, Sam Harding, Angus McNair, Julie Clayton, Winsome Barrett-Muir, Alan Richardson, Jemima Dooley, Joseph Webb, Tomas Lemke, Elizabeth Coulthard, Nicholas Turner

## Abstract

**INTRODUCTION:** Meaningful research into mild cognitive impairment (MCI) is limited by trial outcomes which are heterogeneous and may not always be important to patients. We developed a core outcome set (COS) for the evaluation of interventions in patients with MCI (MCI-COS).

**METHODS:** A scoping umbrella review and interviews with stakeholders (patients, family members, and professionals) determined a longlist of potential outcomes. A modified two-round Delphi study and consensus meeting agreed the final MCI-COS.

**RESULTS:** A ten-item COS was identified: cognitive functioning (non-memory), memory, mental health and wellbeing, social functioning/relationships, quality of life, everyday functioning and independence, biomarkers of brain health, progression to dementia, general/physical health, and sleep.

**DISCUSSION:** Embedding the COS into clinical trials and practice will reduce outcome heterogeneity and encourage transparent reporting of outcomes prioritised by stakeholders, beyond those typically included in trials for MCI.

## 1 Background

A key strategy for slowing or reversing cognitive impairment and neurodegeneration is to intervene as early as possible [1]. Mild cognitive impairment (MCI), where cognitive decline is evident but functional abilities remain largely preserved [2], is the earliest stage of cognitive impairment routinely diagnosed in clinical practice and recognised as an optimal therapeutic window for disease-modifying treatments for Alzheimer’s disease (AD) [3].

Although historically conceptualised as an intermediate stage between normal cognition and AD dementia [4], MCI is clinically and neuropathologically heterogenous [5]. Patients with MCI may not have AD pathology [6] or have several co-pathologies alongside AD, including Lewy bodies or cerebral infarcts [7].

Clinically, MCI may present with one or multiple cognitive domains affected and whilst memory is often affected (amnestic MCI), memory may also be preserved (non-amnestic MCI) [8]. Prognosis is also highly variable. Patients with MCI have an increased risk of developing dementia but may experience a slow or rapid progression to dementia, reversion to normal cognition, or stable MCI [9]. Whilst some patients with MCI due to AD now have access to anti-amyloid disease-modifying treatments, there remains no effective or appropriate treatment for most patients living with MCI [10–12].

International guidelines and care pathways for MCI vary considerably [2, 13] and focus largely on delaying or preventing dementia onset [2]. There also remains a lack of consensus on which patient outcomes should be prioritised as therapeutic targets, which limits research impact of clinical trials in MCI [14] [15] [16]. Uncertainty around prognosis and the increased likelihood of dementia is understandably often a source of anxiety in patients with MCI [17]. However, other outcomes beyond reducing dementia risk are also important to people living with MCI [18, 19]. Whilst cognitive symptoms are mild, people with MCI often report lower quality of life [20, 21], more neuropsychiatric symptoms, and difficulties maintaining social networks [22, 21, 23, 24]. MCI is also associated with higher healthcare utilisation costs and support needs [25] [26, 27]. To ensure we develop treatments and care pathways which meaningfully help people with MCI, it is crucial to determine what outcomes are priorities both to patients and those who care for them.

A core outcome set (COS) for MCI fills this gap in knowledge. A COS is a standardised minimum agreed set of outcomes for clinical trials in a particular condition [28], designed to ensure that researchers measure and report outcomes most likely to be relevant to stakeholders. Beyond research, COS can also be implemented for patient monitoring in clinical care and outcome selection in systematic reviews [29]. Whilst several COS exist for people living with dementia [27, 30, 31], uncertainty in prognosis coupled with milder symptoms and largely preserved daily functioning likely mean that priorities differ between people living with dementia compared to people living with MCI.

### 1.1 Objectives and scope

The aim of this study was to develop a COS for use in interventional trials and clinical practice to assess the effectiveness of treatments for community-dwelling adults living with MCI, through consultation with stakeholders with lived and professional experience of MCI.

## 2 Methods

### 2.1 Study overview

The COS was developed in five stages: (1) a scoping umbrella review [16]; (2) qualitative interviews with stakeholders; (3) evidence synthesis to create a longlist of potential COS outcomes; (4) a two-round Delphi study; (5) a hybrid consensus meeting to agree the final COS (**Figure 1**).

**Figure 1.**
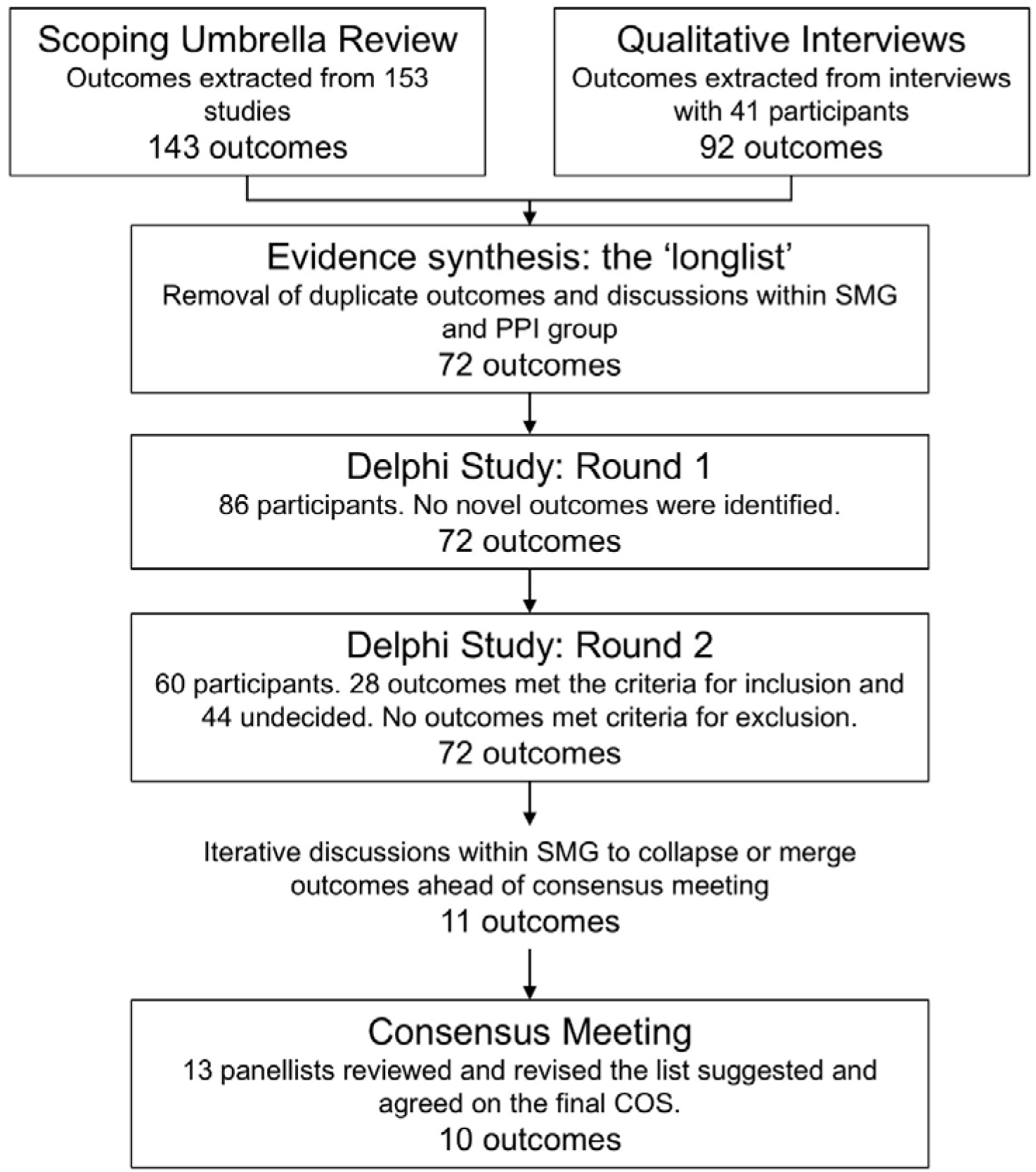
Flow chart summarising the development of a core outcome set for mild cognitive impairment (MCI-COS).

The study protocol was developed using the Core Outcome Measures in Effectiveness Trials (COMET) criteria, prospectively registered with the COMET initiative (registration ID: 2117), and published prior to data analysis [14]. The study ran from June 2023 to July 2025 and is reported in line with the Core Outcome Set-Standards for Reporting (COS-STAR) Statement [32] (Error! Reference source not found.). Each stage of the study was overseen by a multidisciplinary study management group (SMG) consisting of researchers, healthcare professionals, and patient and public involvement (PPI) representatives who met regularly throughout the study period.

### 2.2 Information sources

#### 2.2.1 Stage 1: Scoping umbrella review of interventional studies

An extensive list of potential outcomes was extracted from a scoping umbrella review of interventional trials and is described in detail elsewhere [16].

#### Stage 2: Qualitative interviews

Stakeholders (patients with MCI, family members of individuals with MCI, and professionals who work with individuals with MCI) were invited to participate in semi-structured interviews. Interviews were conducted one-to-one with a researcher in a setting chosen by the participant, either online via videoconferencing, by telephone, or in person. An interview topic guide was developed in collaboration with our patient and public involvement (PPI) group consisting of individuals with lived experience of MCI and dementia to ensure that questions, interview style, and approach were accessible, inclusive, and encouraged open discussions (Error! Reference source not found.).

Interviews were audio-recorded, transcribed, and coded using content analysis to identify outcomes. Outcomes were recorded in an interview codebook.

#### 2.2.2 Stage 3: Evidence synthesis

Outcomes identified in Stage 1 and 2 were iteratively combined and refined through consultations with the PPI group and the SMG. Outcomes were then organised conceptually into outcome domains informed by Dodd’s outcome taxonomy [33].

#### 2.2.3 Stage 4: Delphi surveys

To refine the long-list of outcomes into a COS, a consensus process involving a two-round modified Delphi study was undertaken. Delphi studies, in which a series of ‘rounds’ in which experts are asked their opinion on a particular issue, are often utilised in COS development [34]. Anonymity of individual responses helps to reduce bias, whilst reviewing group and individual responses helps with reflection and movement towards consensus [35].

The traditional Delphi method involves open-ended questions refined subsequently through rounds until consensus is reached [36]. The scoping umbrella review and interviews undertaken prior to the Delphi surveys identified a list of outcomes for consideration based on what is typically measured in clinical trials and what outcomes matter to stakeholders. Therefore, this study used a modified Delphi study to reduce participant burden, limited *a priori* to two rounds with a final consensus meeting [37].

Lay outcome names, definitions, and examples were provided with medical terms (e.g., ‘instrumental activities of daily living’) alongside and were informed by discussions with the PPI group and stakeholder interviews. Participants could opt to complete the Delphi surveys online or on paper. Online Delphi surveys were administered via the REDCap electronic data capture system hosted by the University of Bristol [38].

##### 2.2.3.1 Delphi round one

Round one of the Delphi survey was open from March 10^th^, 2025 to May 8^th^, 2025. Participants were asked to score each of the outcomes on a Likert-scale from one to nine (1-3 = not essential, 4-6 = important but not essential, 7-9 = essential).

Participants were also provided an opportunity to suggest additional outcomes at the end of round one. No outcomes were removed at the end of round one, irrespective of scores.

##### 2.2.3.2 Delphi round two

Round two of the Delphi survey was open from May 15^th^, 2025 to June 10^th^, 2025. Participants who scored any item during round one were invited to participate in round two. Participants were asked to score the same outcomes in the same order as round one. Each outcome was presented for rating alongside the participant’s score from round one and the median scores from each stakeholder group [39].

Participants were reminded that they were not required to change their scoring from the first round.

Consensus was defined *a priori* [14] as ≥70% agreement. In the Delphi study, consensus was considered within each stakeholder group. Outcomes scored as ≥6 by at least 70% participants and ≤2 by less than 10% participants were included in the following stage of the COS development and outcomes scored ≤4 by at least 70% participants and ≤8 by less than 10% participants were excluded. Remaining outcomes were considered ‘undetermined’ and discussed at the consensus meeting.

#### 2.2.4 Stage 5: Consensus meeting

A hybrid consensus meeting was held on June 19^th^, 2025, to discuss the results from the two Delphi surveys and agree the final COS. Participants from different stakeholder groups were invited to participate as a single panel and attend face-to-face in Bristol, United Kingdom or join remotely via videoconferencing. Participants were not asked to disclose their stakeholder group but were welcome to if they wanted to share their experiences. The consensus panel size was not pre-specified and was determined by the willingness and availability of eligible participants to join the consensus meeting.

At the start of the meeting, panel members were provided with a summary of the rationale behind the study and the consensus process, including definitions of key terminology. A list of proposed outcomes was then presented to the panel alongside its median rating from round two of the Delphi survey. Participants were encouraged to discuss each outcome, with ad-hoc votes held when participants suggested renaming or re-grouping outcomes (i.e., collapsing or splitting outcomes). Outcomes that had reached consensus for inclusion through the second round of the Delphi survey were eligible for merging into a collapsed outcome where outcomes were considered too numerous or overlapping.

During the consensus meeting, panel members voted outcomes ‘in’ or ‘out’. Consensus during the final consensus meeting was considered across the entire panel rather than across individual stakeholder groups. Outcomes reaching ≥70% agreement during the consensus meeting were included in the final COS, either within merged or separate outcomes. Finally, participants were asked to vote on the final COS, with consensus defined *a priori* as ≥70% in support of the final COS.

### 2.3 Participants

All participants involved in the consensus process were community-dwelling adults aged 18 or over with capacity to consent. Three stakeholder groups were invited to participate in the study:

1. individuals diagnosed with MCI (‘Patients’). Patients were recruited from NHS cognitive disorders or memory clinics from the South West and North West of

England and research volunteer databases. MCI diagnoses were confirmed via documentation in medical records in the last 12 months.

1. individuals who support someone with MCI in a personal capacity (‘Family members’). Family members were identified through research volunteer databases, word-of-mouth, and community organisations.
2. individuals who support people with MCI in a healthcare, research, or other professional capacity (‘Professionals’). Professionals were identified through professional networks and word-of-mouth. In addition to researchers and healthcare professionals working within primary, secondary, and tertiary care, individuals from non-profit and community organisations, paid agencies which provide care and support for individuals with MCI, and organisations involved in providing guidance for health and social care and relevant policy were eligible to participate.

Where participants met criteria for more than one stakeholder group, they self-selected their stakeholder group but were welcome to share perspectives across their lived and professional experience.

Participants could participate in the qualitative interviews, Delphi surveys, or both. Participants who completed the qualitative interviews were invited to complete the Delphi surveys and participants who completed both rounds of the Delphi study were invited to participate in the final consensus meeting.

### 2.4 Ethics and consent

Ethical approval was received from the London—Queen Square Research Ethics Committee (Reference 23/PR/1580) for the interviews, Delphi study, and consensus meeting. Written informed consent was received from participants prior to engaging in any study activities. The study was conducted in accordance with Good Clinical Practice and the Declaration of Helsinki.

Participants were offered reimbursement for travel expenses incurred during the study. Participants who completed interviews or both Delphi surveys in the Patients and Family members group (participating outside of a professional capacity) were offered gift vouchers as compensation for their time and expertise.

### 2.5 Patient and public involvement (PPI)

Members of the public with lived experience of MCI and dementia (patients, family members, and caregivers) were involved throughout the study to enhance its relevance, inclusivity, transparency, and quality. Two PPI representatives were an integral part of the SMG and contributed to design, delivery, and decision-making throughout the study. Additional PPI contributors supported the design and pilot testing of participant-facing materials, provision of plain English outcomes, and decision-making around data collection methods and outcome merging. Detailed information on PPI is reported in accordance with the revised Guidance for Reporting Involvement of Patients and the Public – Short Form (GRIPP-2) [40] in Error!

Reference source not found.. PPI contributors were paid for their time in line with National Institute for Health and Care Research (NIHR) guidance and reimbursed for any travel expenses incurred.

## 3 Results

Development of the COS is outlined in **Figure 1**. Participant demographics for the qualitative interviews, Delphi surveys, and consensus meeting are summarised in Error! Reference source not found..

### 3.1 Stage 1: Scoping umbrella review

The full results of the review are published elsewhere [16]. Briefly, 143 unique outcomes were generated from 153 studies during the review. Global cognition was the most commonly reported outcome and reported in more than 50% of studies included, with two-thirds of outcomes identified reported by three or fewer studies.

### 3.2 Stage 2: Qualitative interviews

Of the 42 participants who consented to participate in stakeholder interviews, 41 participants completed interviews and one participant was lost to follow-up. Over half of the participants for the stakeholder interviews were patients with MCI (22/41, 53.7%) (**Supplementary Materials D**). Family members were predominantly partners or spouses (5/8, 62.5%). Professional backgrounds included neurology, old age psychiatry, allied health, general practice, and older adult community services.

Ninety-two unique outcomes were generated from the interviews, spanning all five areas of Dodd’s taxonomy (**Supplementary Materials B**). Memory, episodic memory, maintaining independence (IADLs), driving, having meaningful relationships, and participation in leisure or meaningful activities were outcomes reported by the highest number of participants and were mentioned by participants across all three stakeholder groups.

### 3.3 Stage 3: Evidence synthesis (creating the ‘longlist’)

Outcomes identified during Stage 1 and 2 were iteratively merged and refined through discussion with PPI representatives in online and face-to-face groups, until the SMG agreed upon 72 unique outcomes which could be rated in the Delphi surveys (**Supplementary Materials C**). Some outcomes were collapsed under broader outcome names (e.g., planning was collapsed into executive functioning), and similar outcomes were grouped together (e.g., self-confidence and self-efficacy). One outcome identified during the review, deqi sensations, was removed by the SMG as it is an outcome with specific relevance to a single intervention (acupuncture).

Most outcomes were identified during both the scoping umbrella review and qualitative interviews (56/72, 77.8%). Six outcomes were based solely on outcomes identified during the review: telomere length; circadian rhythms; markers of cardiorespiratory health; psycho-motor skills; health-related quality of life; and working memory. Ten outcomes were identified solely during the qualitative interviews: survival; action-based memory (procedural memory); being able to find your way around (navigation); feeling frustrated; sense of agency, autonomy, or control; feeling stigmatised; self-identity; ability to drive; feeling tired or sleepy during the day; treatment costs.

### 3.4 Stage 4: Delphi surveys

Of the 103 participants who consented to participate in the Delphi study, 86 participants rated at least one outcome in round one and 60 participants rated at least one outcome in round two, giving a retention rate between Delphi survey rounds of 69.8%. In both rounds, there was representation from all stakeholder groups with an approximately even split between participants with lived experience (Patients and Family members) and Professionals. Demographic information is provided in detail in Error! Reference source not found..

No new outcomes were added at the end of round one. However, changes were made to definitions to incorporate suggestions made by participants during round one and to simplify the instructions.

At the end of round two, 28 outcomes met *a priori* criteria for inclusion, and 44 outcomes met our criteria for ‘undetermined’, requiring further consideration at the consensus meeting. No outcomes met the criteria for exclusion.

Patients rated outcomes highly overall, with only two outcomes (2.8%) receiving a median score of ≤ 6 in round one and four outcomes (5.6%) receiving a median score of ≤ 6 in round two. In contrast, Family members provided median ratings ≤6 for 33 (45.8%) and 34 (47.2%) outcomes, and Professionals provided median ratings ≤6 for 36 (50.0%) and 37 (51.4%) outcomes in round one and round two, respectively (Error! Reference source not found.).

In round two, overall cognitive functioning and time to progression to dementia ranked within the top five outcomes across all three stakeholder groups. Time to progression to dementia ranked highest among patients, overall cognitive functioning ranked highest among family members, and maintaining independence (IADLs) ranked highest among professionals.

### 3.5 Stage 5: Consensus meeting

As no outcomes were dropped during the Delphi study, and 28 outcomes had been voted ‘in’, the SMG met to discuss how to manage the consensus meeting and final COS.

Several outcomes were grouped based on similarity into broader outcomes more closely resembling the granularity of outcome domains in Dodd’s outcome taxonomy [41] (**Supplementary Materials E**). For example, overall cognitive functioning was combined with 9 additional outcomes relating to more specific cognitive domains to become a single outcome of cognitive functioning. Two outcomes (possible side effects and satisfaction and engagement with the treatment) were suggested to be removed as these were not effectiveness outcomes and instead more closely reflected good practice for trial reporting without specific relevance to MCI.

An initial list of 11 effectiveness outcomes covering life impact (cognitive functioning, emotional functioning/wellbeing, social functioning/relationships, global quality of life, health-related quality of life, independence/functional ability) and physiological/clinical outcomes (psychiatric outcomes, biomarkers of brain health, (likelihood of) progression to dementia, general/physical health, sleep) was suggested as a starting point for the consensus panel discussions, though participants were encouraged to suggest alternatives and improvements.

Participants were also presented with the two non-effectiveness outcomes (possible side effects and satisfaction and engagement with treatment) and asked whether they would consider them core outcomes relating to effectiveness.

All participants who indicated interest in and availability for the consensus meeting were invited to participate. Fifteen participants agreed to join the consensus meeting. Two participants (both Patients) were unable to attend on the day due to personal circumstances, resulting in 13 experts on the final consensus panel. Seven joined remotely and six attended in person. There was an approximately even split between all three stakeholder groups (**Supplementary Materials D**).

During the consensus meeting, several changes to the proposed list were agreed upon to create the final COS. Cognitive functioning was split into two outcomes: memory and cognitive functioning (non-memory). Emotional functioning/wellbeing and psychiatric outcomes were merged into a single outcome (mental health and wellbeing) as were global quality of life and health-related quality of life (global quality of life). Two outcomes were re-named: independence/functional ability was renamed to everyday functioning and independence and (likelihood of) progression to dementia was shortened to progression to dementia. The non-effectiveness outcomes (possible side effects/adverse events and satisfaction and engagement with treatment) were considered important to report as standard across clinical trials but not considered as core outcomes for MCI specifically. The remaining outcomes suggested by the SMG were agreed upon without changes.

From the 44 outcomes which did not achieve ‘consensus in’ and were undetermined, participants put forward 9 outcomes for panel discussion and voting during the consensus meeting. Eight outcomes were voted out (**Supplementary Materials E**). One outcome, memory of experiences and events (episodic memory), was voted in but merged with the existing memory outcome.

#### 3.5.1 Final COS

The final COS included ten outcomes recommended to be measured in all interventional trials for MCI: cognitive functioning (non-memory), memory, mental health and wellbeing, social functioning/relationships, global quality of life, everyday functioning and independence, biomarkers of brain health, progression to dementia, general/ physical health, and sleep (**Table 1**. Final core outcome set for mild cognitive impairment (MCI-COS). All outcomes were agreed with 100% consensus during the final consensus meeting.**Table 1** and **Figure 2**).

**Figure 2.**
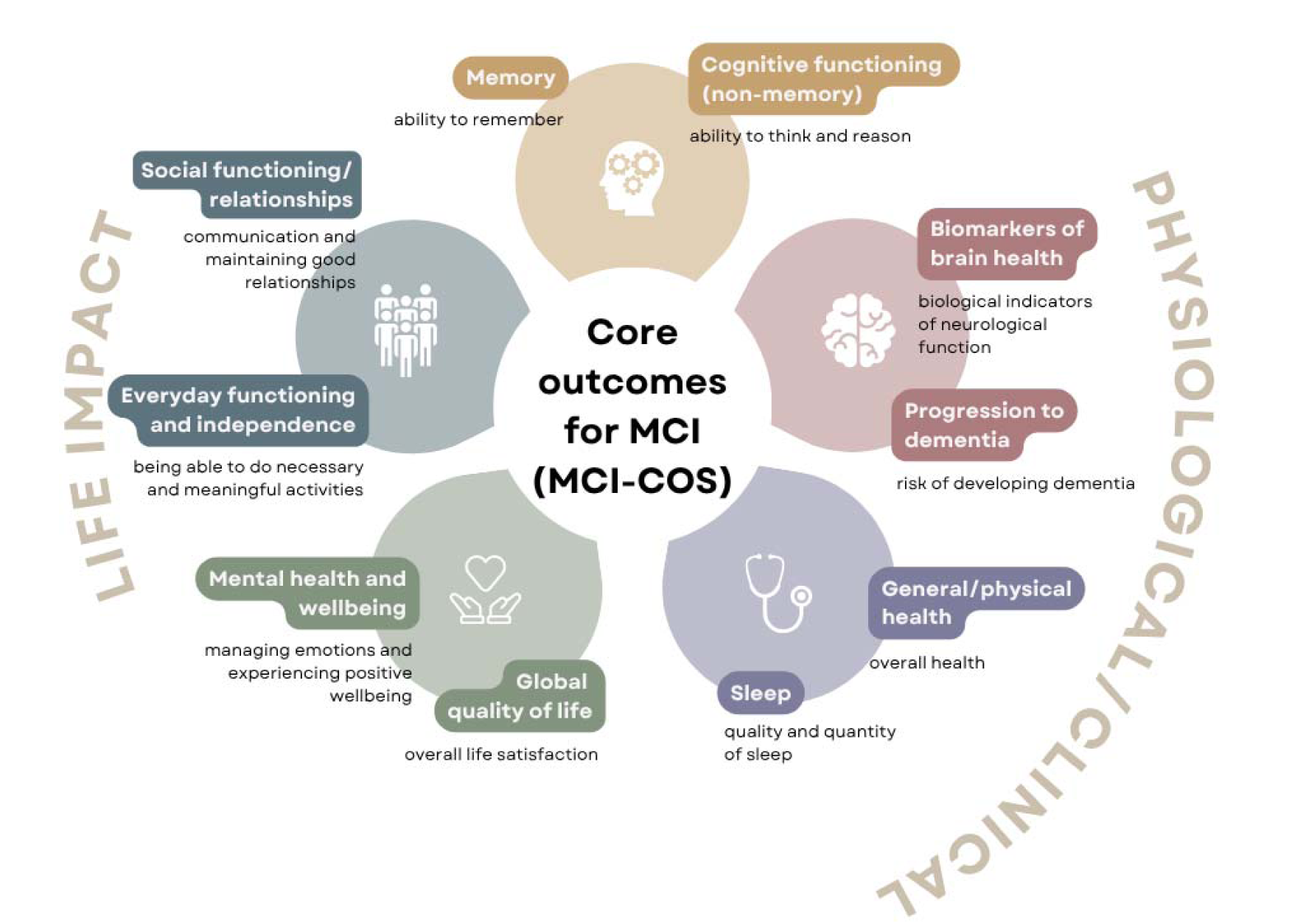
Final core outcome set for mild cognitive impairment (MCI-COS).

**Table 1.**
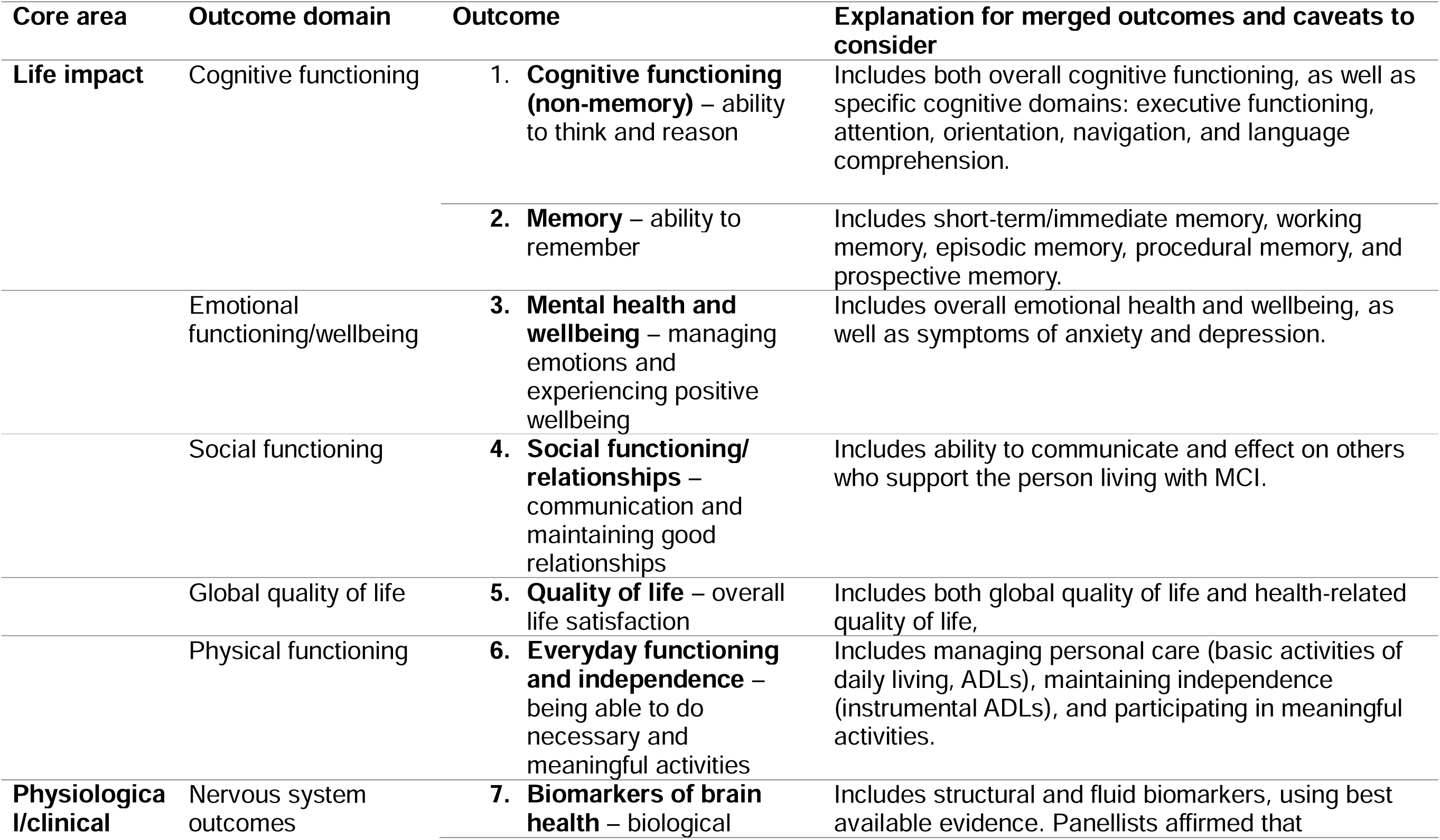
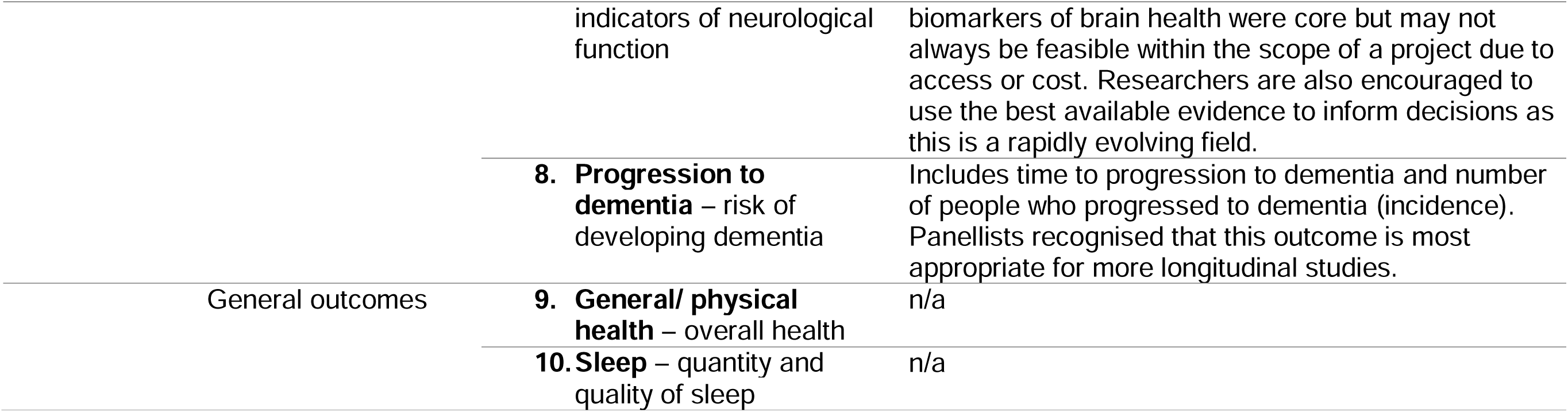
Final core outcome set for mild cognitive impairment (MCI-COS). All outcomes were agreed with 100% consensus during the final consensus meeting.

Eleven panellists voted on the final COS, with all panellists supporting the final COS.

## 4 Discussion

Here, we present a core outcome set for MCI (the MCI-COS): ten outcomes identified as ‘core’ or priority outcomes for understanding the effectiveness of interventions for people living with MCI. In line with COMET guidance [34], stakeholders including patients, family members, healthcare professionals, and researchers shaped the COS throughout its development through interviews, surveys, a final consensus meeting, and extensive patient and public involvement. Uptake of the MCI-COS will support trials, systematic reviews, and clinical practice to capture and report patient-centred outcomes and help determine whether treatments provide a meaningful benefit for people living with MCI.

The MCI-COS includes outcomes currently often reported by trials for MCI and prodromal AD such as cognition, everyday functioning, and biomarkers [16].

Determining meaningful clinical benefit in cognitive and functional outcomes in patients with MCI has long been debated []. However, there has been progress with establishing minimally clinically important differences, the smallest unit of change perceived as a clinically meaningful for patients with MCI, for key outcome measures including the Mini-Mental State Examination (MMSE) and integrated Alzheimer’s Disease Rating Scale (iADRS) [42] [42]. Biomarkers can offer important insights into disease progression and therapeutic action, but it is less clear whether changes reflect clinical benefit and as such biomarkers are often included as secondary trial outcomes . Beyond these frequently used measures, the MCI-COS also includes broader physiological and life impact outcomes that are infrequently reported in trials, reflecting an important gap between the priorities of stakeholders and trial design. For example, in our review of 153 interventional studies enrolling participants with MCI, only one reported an outcome relating to sleep quality and five studies included outcomes relating to social functioning and relationships . The importance of multidimensional patient-reported, quality of life, and neuropsychiatric outcomes, beyond core disease outcomes, for determining meaningful patient benefit is increasingly recognised and will help to support the translation of treatments from research into clinical practice [19, 43]. Through centring outcome reporting and trial design around what matters most to patients and those involved in their care, the MCI-COS provides a framework for trials to move towards a more patient-centred and comprehensive approach to outcome reporting and trial design. [19]

It is important to emphasise that outcomes in the COS are recommended as a minimum, and it is expected that additional outcomes will be added [44]. Our COS was developed for relevance to MCI of any underlying aetiology and across intervention types. When designing trials for patients with MCI, we recommend trialists consider whether including outcomes from disease-specific COS (e.g., for AD [45] or Parkinson’s disease [46]) or intervention-specific COS (e.g., nutrition [47] or sleep [48]) may add further value. It is also recommended that regulatory agencies are consulted to advise on specific outcomes that may be required for adoption into practice. In the United States, the Food and Drug Administration currently recommend cognition, pathophysiology, functional change, and/or delaying progression to dementia as outcomes in trials for patients with MCI due to AD [49, 50], which aligns closely with our COS.

Whilst there is some overlap between the MCI-COS and the 13-item COS for dementia [51] and the recently developed *What Matters Most™* (WMM) conceptual model for AD [52], there was greater breadth of outcome domains included in the MCI-COS. Both the dementia COS and the WMM focused on life impact outcomes, whilst the MCI-COS is broader and specifically identifies four clinical/physiological outcomes, including outcomes both specific to MCI (e.g., progression to dementia) and more general outcomes (e.g., sleep). Such differences may reflect the more uncertain prognosis for MCI, compared to dementia and or having biomarkers of AD, or how outcomes of importance may differ depending on the stage of cognitive impairment [18, 53]. However, both COSs and WMM highlight a core common theme among individuals living with cognitive impairment and neurodegeneration: the importance of capturing life impact outcomes beyond cognition and into social, emotional, and everyday functioning.

Excessively large COS are not feasible to implement due to time and cost burden, whilst missing key outcomes considered meaningful or relevant to patients may also threaten participant recruitment and retention [54]. Whilst it is possible that a 10-item COS may result in recommendations for 10 or more different measures, it is also possible for the COS to be covered by fewer, broader outcome measurement instruments. For example, multidimensional assessments such as the WHOQOL-BREF [55] or the 36-item Short Form Health Survey (SF-36) [56] cover multiple outcomes from the MCI-COS within a single patient-reported outcome measurement instrument. Development of a core measurement set will be needed to ensure consistent outcome reporting and identify measures which meaningfully capture the COS with minimal burden.

Outcome granularity is a recognised challenge in COS development [57]. Previous research has suggested that physiological/clinical outcomes are generally specified at a more granular level than life impact outcomes [33]. During our consensus process, physiological/clinical outcomes were merged and in the final COS reflected similar granularity to life impact outcomes. Interestingly, the most frequently used outcome identified in our review [16], global cognition, was considered too broad to be a meaningful outcome during our consensus process. Both consensus panellists and PPI discussions emphasised the importance of capturing and separately reporting both memory and non-memory cognitive outcomes, which maps to the amnestic and non-amnestic cognitive subtypes often used to delineate MCI patients [58].

### 4.1 Limitations

Previous Delphi studies have identified difficulty achieving consensus through the Delphi study methodology [59, 60]. Ratings across all outcomes in the Delphi study were high, with 28 outcomes reaching our a priori cut-off for inclusion in the final COS and no outcomes being voted out. Patients rated nearly all outcomes as essential to include, highlighting good content validity of the longlist and its relevance to patients. However, it was necessary to collapse more granular outcomes to align more closely to the granularity of outcome domains [41] during the consensus process and collapsing outcomes at an earlier stage may have reduced participant burden. As patient burden and costs associated with measuring large COS are potential barriers to COS uptake [44], we collapsed similar outcomes to result in a final COS consisting of ten outcomes. Whilst reaching unanimous agreement among the consensus panellists, it is possible that these merged or collapsed outcomes are less reflective of the breadth of outcomes considered core by the Delphi study participants. Our findings may also have been influenced by methodological decisions made *a priori.* The use of a nine-point rating scale, although commonly utilised in COS development [34], tends to result in a higher number of outcomes being rated as essential compared to a three-point scale [61]. We also used the criteria for consensus as defined by Williamson and colleagues [62], and more stringent criteria for consensus could also have resulted in a smaller COS but equally could have missed important outcomes [63].

Additionally, outcomes more closely relating to health economics, safety, and feasibility were identified during the review and interviews and included as potential outcomes in the long list for the Delphi study. Two of these non-effectiveness outcomes, possible side effects and satisfaction and engagement with treatment, were rated ‘in’ as outcomes during the Delphi study. However, the SMG and consensus panel both agreed that whilst important, these were not outcomes relating to treatment effectiveness and would be expected to be reported as standard, without specific relevance to MCI. Therefore, these outcomes should have been dropped at an earlier stage and are not included in the final MCI-COS, but their importance is highlighted.

Although efforts were made to recruit a diverse panel of stakeholders, certain regions and professional groups were underrepresented, including those from low-and middle-income countries and stakeholders involved in policy and regulatory approvals, which may affect international generalisability and COS uptake.

Finally, it was beyond the scope of this work to identify the most appropriate outcome measure for each item in the COS. The consensus meeting highlighted the importance of considering the practicalities of measuring physiological/clinical outcomes in research and clinical practice, including access, costs, and how best practice evolves over time. For example, whilst imaging and cerebrospinal fluid biomarkers of neurodegeneration are more established, blood-based biomarkers are increasingly utilised in research and may offer a more accessible alternative [64]. Some outcomes may also be more difficult than others to demonstrate a meaningful improvement, particularly at shorter timeframes. Continued discussions around measuring everyday functioning and cognition in MCI [65], the use of surrogate endpoints such as fluid biomarkers where changes may be more easily observable [49], and what constitutes clinically meaningful benefits [42] are needed. To support widescale and consistent implementation of the COS, further work is needed to develop a core measurement set based on the MCI-COS [66, 44], as well as establish what could be considered a minimal clinically important difference for each of these outcome measures [67].

### 4.2 Conclusions

Despite affecting over one in five community-dwelling older adults [23, 24, 25], there are currently no disease-modifying treatments for majority of MCI patients worldwide and effective pharmacological and non-pharmacological treatments are needed. To reduce research waste and accelerate research into effective treatments, we have developed a COS relevant to people with lived experience of MCI, clinicians, and researchers. The MCI-COS could help to ensure meaningful outcomes are consistently and transparently reported and support future synthesis of research across studies – but only if trialists decide to use it.

## Data Availability

All data produced in the present study are available upon reasonable request to the authors.

## 6 Acknowledgements

We would first and foremost like to thank all participants who took part in the research, the family members who supported them, and the many public contributors from the ReMemBr Lived Experience Experts Group who helped to shape and support the research throughout the process. We would also like to thank the research teams at participating sites: North Bristol NHS Trust, Avon and Wiltshire Mental Health Partnership NHS Trust, Greater Manchester Mental Health NHS Foundation Trust, and Cornwall Partnership NHS Foundation Trust.

## 6.1 Statement of Contribution

**VGG:** Conceptualisation, Data curation, Formal analysis, Investigation, Methodology, Project administration, Resources, Software, Validation, Visualisation, Writing – Original draft. **SH:** Conceptualisation, Methodology, Supervision, Writing – Review & editing. **AGKN:** Conceptualisation, Funding acquisition, Methodology, Supervision, Writing – Review & editing. **JC:** Funding acquisition, Resources, Supervision, Writing – Review & editing. **WBM:** Funding acquisition, Supervision, Writing – Review & editing. AR: Funding acquisition, Supervision, Writing – Review & editing. **JD:** Funding acquisition, Resources, Writing – Review & editing. **JW:** Funding acquisition, Resources, Writing – Review & editing. **TL:** Data curation, Investigation, Formal analysis, Writing – Review & editing. **EC:** Conceptualisation, Funding acquisition, Methodology, Supervision, Writing – Review & editing. **NT:** Conceptualisation, Formal analysis, Funding acquisition, Methodology, Supervision, Writing – Review & editing.

## 6.2 Funding Sources

This research was funded by the National Institute for Health and Care Research (NIHR) Research for Patient Benefit programme (NIHR204135). The views expressed are those of the authors and not necessarily those of the NIHR or the Department of Health and Social Care.

## 6.3 Conflicts of Interest

Whilst not considered a conflict of interest, for full disclosure EC has received funding for education and consultancy at Eisei, Biogen, Lilly, and AstronautX. No other authors declare a conflict of interest.

## Notes

### Author Declarations

NHS research ethics committee of London-Queen Square Research Ethics Committee gave ethical approval for this work (Reference 23/PR/1580).

