## Supplementary Files for "Development of a Core Outcome Set for Mild Cognitive Impairment (MCI-COS): Recommendations from a multistakeholder Delphi consensus study"

### Supplementary Materials A

##### Core Outcome Set-STandards for Reporting: The COS-STAR Statement Checklist

| **SECTION/TOPIC** | **ITEM No.** | **CHECKLIST ITEM** | **RELEVANT SECTION(S)** |
| --- | --- | --- | --- |
| TITLE/ABSTRACT | | | |
| Title | 1a | Identify in the title that the paper reports the development of a COS | Title page |
| Abstract | 1b | Provide a structured summary | Abstract |
| INTRODUCTION | | | |
| Background and Objectives | 2a | Describe the background and explain the rationale for developing the COS. | Background |
|  | 2b | Describe the specific objectives with reference to developing a COS. | Objectives and scope |
| Scope | 3a | Describe the health condition(s) and population(s) covered by the COS. | Objectives and scope |
|  | 3b | Describe the intervention(s) covered by the COS. | Objectives and scope |
|  | 3c | Describe the setting(s) in which the COS is to be applied. | Objectives and scope |
| METHODS | | | |
| Protocol/Registry Entry | 4 | Indicate where the COS development protocol can be accessed, if available, and/or the study registration details. | Study overview |
| Participants | 5 | Describe the rationale for stakeholder groups involved in the COS development process, eligibility criteria for participants from each group, and a description of how the individuals involved were identified. | Participants |
| Information Sources | 6a | Describe the information sources used to identify an initial list of outcomes. | Information sources |
|  | 6b | Describe how outcomes were dropped/combined, with reasons (if applicable). | Results |
| Consensus Process | 7 | Describe how the consensus process was undertaken. | Information sources |
| Outcome Scoring | 8 | Describe how outcomes were scored and how scores were summarised. | Information sources |
| Consensus Definition | 9a | Describe the consensus definition. | Methods |
|  | 9b | Describe the procedure for determining how outcomes were included or excluded from consideration during the consensus process. | Methods, Results |
| Ethics and Consent | 10 | Provide a statement regarding the ethics and consent issues for the study. | Ethics and consent |
| RESULTS | | | |
| Protocol Deviations | 11 | Describe any changes from the protocol (if applicable), with reasons, and describe what impact these changes have on the results. | Information sources, Results |
| Participants | 12 | Present data on the number and relevant characteristics of the people involved at all stages of COS development. | Results, Supplementary Materials |
| Outcomes | 13a | List all outcomes considered at the start of the consensus process. | Results, Supplementary Materials |
|  | 13b | Describe any new outcomes introduced and any outcomes dropped, with reasons, during the consensus process. | Results |
| COS | 14 | List the outcomes in the final COS. | Results |
| DISCUSSION | | | |
| Limitations | 15 | Discuss any limitations in the COS development process. | Discussion |
| Conclusions | 16 | Provide an interpretation of the final COS in the context of other evidence, and implications for future research. | Discussion |
| OTHER INFORMATION | | | |
| Funding | 17 | Describe sources of funding/role of funders. | Funding sources |
| Conflicts of Interest | 18 | Describe any conflicts of interest within the study team and how these were managed. | Conflicts of interest |

From: Kirkham JJ, Gorst S, Altman DG, Blazeby JM, Clarke M, Devane D, et al. (2016) Core Outcome Set–STAndards for Reporting: The COS-STAR Statement. PLoS Med 13(10): e1002148. https://doi.org/10.1371/journal.pmed.1002148

### Supplementary Materials B

##### Topic guide for Qualitative Interviews

Below is the Topic Guide for interviews with patients. Similar questions were used for all stakeholder groups, with minor adjustments to wording.

###### Aims and Objectives

- To explore patients’ experiences of living with MCI.
- To determine what the patient feels are the most important outcomes for research into MCI.
- To determine the most suitable instruments for MCI research.
- To determine appropriate language to describe outcomes, to inform development the Delphi process.

###### Introduction

- Is today a good day to go ahead with the interview or would you prefer to reschedule?
- Interviewer Name and background
- Explain expected aims, sensitive subjects and duration of interview and confidentiality.
- Confirm consent to qualitative interview and recording.
- How this research will benefit others, what we hope to achieve, and what their participation could mean for the wellbeing of others

###### History

- How long ago did you receive your MCI diagnosis?
- Who told you your diagnosis? (e.g., memory clinic, GP)
- What were the first symptoms of MCI you noticed?
- What prompted you to go to a doctor?

###### Interview questions

- Does MCI affect your life on a daily basis? If so, in what ways?
  - What does a good day look like for you?
  - What about a day which is not so good?
- What are your strategies for living with your MCI symptoms?
  - How successful are these strategies in managing your symptoms?
  - Are there symptoms you find more challenging to manage yourself?
- Imagine there is something new (e.g., a lifestyle change or a medication) which could help someone with MCI. What improvements would you want this new treatment or change to make to your life?
- Are there aspects of living with MCI that you think doctors and health professionals don’t understand?
- What are your priorities in life right now? What is most important to you?
  - Have these priorities changed since you learned about your MCI diagnosis?

###### Summary and Outcomes Question

- You have said X, Y, and Z are important for you in living well with MCI. Is there anything you want to add?
- Do you think we can measure these things in research? How can we do this? (Give examples, questionnaires, interviews, thinking and memory tests, brain scans, asking family members/close friends)

###### Closing remarks

- Do you have any questions regarding this research?

### Supplementary Materials C

#### GRIPP-2

Patient and public involvement (PPI) in the development of the core outcome set (COS) for mild cognitive impairment (MCI), in line with the revised Guidance for Reporting Involvement of Patients and the Public – Short Form (GRIPP-2).

| Section and topic | Item |
| --- | --- |
| 1. **Aim**   Report the aim of the study | **Study aim**  To develop consensus among key stakeholders – patients, family members, and relevant professionals – on the most meaningful (core) outcomes to collect and report in interventional studies for patients with mild cognitive impairment (MCI).  **PPI aim**  To design a study which is acceptable and inclusive for participants and supports them to provide honest and open answers on what outcomes matter most in MCI. |
| 2. **Methods**   Provide a clear description of the methods used for PPI in the study | Two PPI co-applicants (WBM and AR) were recruited to the study management group and supported all stages in the project from grant application through to dissemination. They attended monthly study management group meetings and advised on recruitment, PPI, and dissemination strategy (including reviewing recruitment for communities often underserved by research, attending PPI meetings, and identifying community organisations who could support recruitment).  Through in-person workshops, online meetings, and email exchanges, lived experience experts (patients with MCI or dementia and family members) advised on information provided to participants prior to and during their involvement in the study, co-developed the stakeholder interview topic guide, guided the wording, interpretation, and definition of outcomes and provided illustrative examples, and piloted and provided feedback on the Delphi surveys. |
| 3. **Results**   Outcomes – Report the results of PPI in the study, including both positive and negative outcomes | Across 10 meetings, 19 lived experience experts contributed to the study in several ways, including:   - Reviewing the study protocol, highlighting the importance of flexibility in data collection (face to face, online, or by telephone; different locations) - Identifying questions that prospective participants may want to know the answers to before taking part, so this information could be provided in the information sheet - Designing questions for the non-professional stakeholder groups which did not involve research jargon, asked about the same topic from different outcomes, and avoided assumptions around experience of MCI to ensure we got the information we needed, and people understood the purpose of the interview - Using positive or neutral wording around outcomes (e.g., executive function, memory), rather than focusing on deficits or difficulties (e.g., executive dysfunction, memory problems) - Advising whether outcomes grouped be grouped under a broader outcome or were considered ‘unique’ - Interpreting and classifying ambiguous outcomes described in stakeholder interviews - Highlighting the importance of clear definitions and giving several examples to illustrate an outcome in the Delphi survey and including researcher jargon outcomes in parentheses, with the lay outcome given first - Identifying that participants may have difficulty deciding whether to answer what they think themselves versus what they think others might feel or want - Piloting the Delphi survey to test whether it worked, how long it took, and if the design was accessible - Advising that participants should be able to return to complete the survey - Supporting dissemination, including reviewing manuscripts and lay summaries |
| 4. **Discussion**   Outcomes – Comment on the extent to which PPI influenced the study overall. Describe positive and negative effects | PPI was highly effective and influenced important aspects of the study. We encouraged input from patients newly diagnosed with MCI who had not previously been a PPI contributor, as well as those who had more experience in PPI. We had a PPI lead and two PPI contributors in our study management group, with extensive experience in PPI who could facilitate meetings and advise on PPI strategy. PPI contributors were given opportunities to be involved throughout the study, and often contributors helped at more than one stage of the research process (e.g., at grant application and co-designing the topic guide). PPI contributors were reimbursed for their time and expertise on the project and supported throughout the process by the PPI lead.  However, there were limitations. At some research stages (e.g., during analysis of the qualitative interviews), we did not identify PPI contributors who were interested or able to contribute, whilst at other stages, the time for feedback was short (e.g., during development of the Delphi survey), which limited the amount of feedback received. At times, conflicting feedback was received (e.g., whether questions were worded appropriately), so it was difficult to integrate all opinions. Finally, we found that in-person meetings, whilst difficult to coordinate and more expensive (e.g., costing for travel expenses), were more productive and PPI contributors were more engaged. In some online meetings, there was more confusion around the purpose of the meeting (e.g., whether they were helping to define/refine outcomes, or sharing what outcomes *they* thought mattered) and it could be difficult to re-focus the meeting. |
| 5. **Reflections**   Critical perspective – Comment critically on the study, reflecting on the things that went well and those that did not, so others can learn from this experience | Lived experience experts helped to significantly shape the study throughout the research life cycle.  Developing a core outcome set can sometimes feel far removed from immediate concerns of patients and family members. However, we felt the input from PPI contributors throughout the study helped to ensure participants with lived experience remained engaged and understood the purpose of each stage of the consensus process.  Two key challenges were identified: the timescales required for feedback and understanding that we were asking to help design the questions and word outcomes, rather than decide on the outcomes to be included in the core outcome set. If the study was repeated, a longer timescale and more in-person workshops or meetings would be beneficial, though this may exclude contributors who live further away or are unable to travel. |

### Supplementary Materials D

#### Full demographics for expert panels involved in the development of the mild cognitive impairment (MCI) core outcome set (COS).

|  | **Qualitative interviews,**  n=41 | **Delphi survey Round one,**  n=86* | **Delphi survey Round two,**  n=60* | **Consensus**  **Meeting,**  n=13 |
| --- | --- | --- | --- | --- |
| Stakeholder group, n (%) | | | | |
| Patients | 22 (53.7%) | 33 (38.4%) | 21 (35.0%) | 5 (38.5%) |
| Unknown aetiology | 10 (24.4%) | 16 (18.6%) | 9 (15.0%) | 2 (15.4%) |
| MCI due to probable AD | 6 (14.6%) | 6 (7.0%) | 6 (10.0%) | 1 (7.7%) |
| MCI due to probable vascular cognitive impairment | 2 (4.9%) | 7 (8.1%) | 3 (5.0%) | 1 (7.7%) |
| MCI due to probable PD or LBD | 1 (2.4%) | 1 (1.2%) | 1 (1.7%) | 1 (7.7%) |
| Other or mixed pathology | 4 (9.8%) | 3 (3.5%) | 2 (3.3%) | 0 |
| Family members | 8 (19.5%) | 11 (12.8%) | 11 (18.3%) | 4 (30.8%) |
| Professionals | 11 (26.8%) | 42 (48.8%) | 28 (46.7%) | 4 (30.8%) |
| Health and social care | 7 (17.1%) | 22 (25.6%) | 15 (25.0%) | 2 (15.4%) |
| Researcher | 3 (7.3%) | 17 (17.1%) | 11 (18.3%) | 2(15.4%) |
| Other service provider (e.g., charity, homecare) | 1 (2.4%) | 3 (3.5%) | 2 (3.3%) | 0 |
| Age at consent (years), n (%) | | | | |
| 29 or under | 0 | 4 (4.7%) | 3 (5.0%) | 0 |
| 30-39 | 6 (14.6%) | 9 (10.5%) | 8 (13.3%) | 2 (15.4%) |
| 40-49 | 7 (17.1%) | 19 (22.1%) | 13 (21.7%) | 2 (15.4%) |
| 50-59 | 2 (4.9%) | 14 (16.3%) | 8 (13.3%) | 2 (15.4%) |
| 60-69 | 9 (22.0%) | 15 (17.4%) | 9 (15.0%) | 1 (7.7%) |
| 70-79 | 12 (29.3%) | 19 (22.1%) | 15 (25.0%) | 5 (38.5%) |
| 80-89 | 5 (12.2%) | 6 (7.0%) | 4 (6.7%) | 1 (7.7%) |
| Gender, n (%) | | | | |
| Male | 19 (46.3%) | 33 (38.4%) | 24 (40.0%) | 6 (46.2%) |
| Female | 22 (53.7%) | 53 (61.6%) | 36 (60.0%) | 7 (53.8%) |
| Ethnicity, n (%) | | | | |
| White | 35 (85.4%) | 74 (86.0%) | 53 (88.3%) | 12 (92.3%) |
| English/Welsh/Scottish/Northern Irish/British | 29 (70.7%) | 61 (70.9%) | 44 (73.3%) | 11 (84.6%) |
| Irish | 2 (4.9%) | 7 (8.1%) | 5 (8.3%) | 0 |
| Any other White background | 4 (9.8%) | 6 (7.0%) | 4 (6.7%) | 1 (7.7%) |
| Black/African/Caribbean/Black British | 3 (7.3%) | 3 (3.5%) | 1 (1.7%) | 0 |
| African | 0 | 1 (1.2%) | 1 (1.7%) | 0 |
| Caribbean | 1 (2.4%) | 2 (2.3%) | 0 | 0 |
| Any other Black background | 2 (4.9%) | 0 | 0 | 0 |
| Mixed/multiple ethnic groups | 3 (7.3%) | 3 (3.5%) | 1 (1.7%) | 1 (7.7%) |
| White and Black African | 2 (4.9%) | 2 (2.3%) | 1 (1.7%) | 1 (7.7%) |
| Any other mixed background | 1 (2.4%) | 2 (2.3%) | 0 | 0 |
| Asian/Asian British  Chinese  Indian | 0  0  0 | 3 (3.5%)  1 (1.2%)  2 (2.3%) | 3 (5.0%)  1 (1.2%)  2 (2.3%) | 0  0  0 |
| Any other ethnic group  Latino | 0  0 | 1 (1.2%)  1 (1.2%) | 1 (1.7%)  1 (1.7%) | 0  0 |
| Information not provided | 2 (4.9%) | 2 (2.3%) | 1 (1.7%) | 0 |
| Highest level of education, n (%) | | | | |
| Primary education | 0 | 0 | 0 | 0 |
| Secondary education | 11 (26.8%) | 14 (16.3%) | 10 (16.7%) | 3 (23.0%) |
| Further education | 5 (12.2%) | 15 (17.4%) | 11 (18.3%) | 3 (23.0%) |
| Higher education | 25 (61.0%) | 57 (66.3%) | 39 (65.0%) | 7 (53.8%) |
| Employment status, n (%) |  |  |  |  |
| Paid employment | 16 (39.0%) | 48 (55.8%) | 33 (55.0%) | 7 (53.8%) |
| Retired | 20 (48.8%) | 32 (37.2%) | 25 (41.7%) | 6 (46.2%) |
| Other^†^ | 5 (12.2%) | 6 (7.0%) | 2 (3.3%) | 0 |

**Notes:** *Individuals who rated at least one outcome prior to the survey closing date and were included in analyses; ^†^Includes individuals who identified as carers, homemakers, those in unpaid employment or volunteering roles, and people unable to work due to health.

### Supplementary Materials E

#### Outcomes identified during qualitative interviews

Each outcome is grouped by Dodd's taxonomy (Core Area, then Outcome Domain) alongside definitions, illustrative quotes, and which stakeholder groups mentioned the outcome. Outcomes identified by all three stakeholder groups are listed first within each outcome domain.

|  | Code | Outcome domain | Working definition | Illustrative quote(s) | Stakeholder group(s) |
| --- | --- | --- | --- | --- | --- |
| Death |  |  |  |  |  |
| 1 | Survival | Mortality/survival | Continuing to live or exist. | "Probably waking up every morning." | Patients |
| Physiological/clinical | |  |  |  |  |
| 2 | Sleep quality | General outcomes | Getting enough sleep and waking feeling that you have slept well or are rested. | "I don't sleep properly or enough really." | Patients, Family members, and Professionals |
| 3 | Symptoms of depression | Psychiatric outcomes | A mental health condition involving prolonged (long-term) low mood or loss of interest in activities you used to enjoy, severe enough to impact daily living. | "I do get down. I do get depressed." | Patients, Family members, and Professionals |
| 4 | Symptoms of anxiety | Psychiatric outcomes | A mental health condition involving prolonger feelings of worry, nervousness, or unease about something with an uncertain outcome, severe enough to impact daily living. | "Sometimes, we have the opportunity to do something, and we won't do it because you would feel anxious." | Patients, Family members, and Professionals |
| 5 | Time to progression to dementia | Nervous system outcomes | Whether the intervention affects the estimated time to convert from MCI to dementia. | "To know how fast or how, how slow it is just to know the progression, timing" | Patients, Family members, and Professionals |
| 6 | Brain health | Nervous system outcomes | How someone's brain and mind thinks, feels, and functions. | "I think maintaining their brain health is a priority." | Patients, Professionals |
| 7 | Eyesight | Physical functioning | Being able to see well. | "I don't watch the television because of my eyes. I'm frightened my eyes are getting worse." | Patients, Family members |
| 8 | Hearing | Physical functioning | Being able to hear well. | "So, I think the hearing affects him a lot because it means he is… you know. So therefore, we've tried to get him a hearing aid." | Patients, Family members |
| 9 | Drinking in moderation | General outcomes | Being able to drink alcohol within recommended limits for good health. | "Not to over booze." | Patients |
| 10 | Having a healthy body weight or BMI | General outcomes | Having a body weight or body mass index within recommended limits for good health. | "Keep my weight down too." | Patients |
| 11 | Eating a healthy diet | Metabolism and nutrition outcomes | Having a balanced diet. | "Try to eat well." | Patients |
| 12 | Brain volume | Nervous system outcomes | The size of the brain's grey and white matter. | "The brain scan I suppose, would at least tell me how much brain I’ve got left." | Patients |
| 13 | Headaches | Nervous system outcomes | Pain or discomfort in your face or head. | "It's like a nauseous feeling. Yeah, headaches. Not really bad headaches, but you know." | Patients |
| 14 | Neuroinflammatory biomarkers | Immune system outcomes | Markers indicating the presence or strength of an immune response. | "You'd have to have a neuroinflammatory component." | Professionals |
| 15 | Nutritional biomarkers | Metabolism and nutrition outcomes | Markers of your metabolic or nutritional health. | "We would be interested in, maybe looking at fat-related biomarkers, like changes in cholesterol et cetera or looking at changes in vitamin C, vitamin D." | Professionals |
| 16 | Biomarkers of pathology/aetiology | Nervous system outcomes | Markers used to assess the risk, presence, or progression of a disease. | "It's biomarkers of amyloid and tau." | Professionals |
| 17 | Presence of risk factors for dementia | Nervous system outcomes | Engaging in behaviours or having certain characteristics which increase the likelihood of developing dementia. | "You can measure the presence of risk factors for dementia and cognitive impairment." | Professionals |
| 18 | Brain activity | Nervous system outcomes | Electrical and chemical activity underlying cognitive and neurological functions. | "A measure of brain activity derived from fMRI EEG/MEG, something like that." | Professionals |
| Life Impact |  |  |  |  |  |
| 19 | Attention | Cognitive functioning | Watching, listening to, or thinking about something whilst tuning out other irrelevant or distracting details. | "I still concentrate [on] what I have to do, how to do it, where to go and all that. I don't, too quick, you know, and I'm making mistakes there, which is bad. Yeah, that is lack of concentration." | Patients, Family members, and Professionals |
| 20 | Cognition | Cognitive functioning | The mental processes that take place in the brain, including thinking, attention, language, learning, memory and perception. | "They're not held back by their cognition, but it is definitely not right." | Patients, Family members, and Professionals |
| 21 | Executive function | Cognitive functioning | A set of cognitive skills that help people manage their thoughts, actions, and emotions to achieve goals. | "Sorting it all out is more difficult." | Patients, Family members, and Professionals |
| 22 | Language comprehension | Cognitive functioning | Being able to understand meaning of spoken or written language. Difficulties in language comprehension are sometimes called Wernicke's (or receptive) aphasia. | "I find it quite difficult understanding what people are saying. Now the first thing that comes to mind is that, you know, you've gone deaf. But that's not true. I can hear it, but I can't understand it." | Patients, Family members, and Professionals |
| 23 | Language production | Cognitive functioning | Being able to speak clearly and “effortlessly”. Difficulties in speaking fluently are sometimes called Broca’s (or expressive) aphasia. | "I think the most important thing is being, is trying to be more fluent during conversations. Rather than being interrupted. Because I've either forgotten my, you know, lost the trace completely, or lost a word." | Patients, Family members, and Professionals |
| 24 | Memory (general) | Cognitive functioning | Being able to remember information, experiences, and people. | "You're not remembering like you're used to remember." | Patients, Family members, and Professionals |
| 25 | Episodic memory | Cognitive functioning | Being able to recall specific past events, along with the details and context of those events. | "I'm just forgetting things, you know, forgetting where I put things and I'm spending half my life looking for something." | Patients, Family members, and Professionals |
| 26 | Problem-solving | Cognitive functioning | Being able to think flexibly, reason abstractly, make decisions by weighing up options, and solve problems. | "If I'm trying to look at finances and money and things like that. When I was asked if I, if I'm trying to decide whether to move something from one from, you know, one account to another or the interest rates and all those sorts of things. I just sort of don't feel as confident as I used to about doing that sort of thing." | Patients, Family members, and Professionals |
| 27 | Subjective cognitive awareness | Cognitive functioning | A person's perception of their cognitive ability, independent of objective standards or performance. | "He knows that he's becoming less tolerant. He knows that he's not finding his words. He knows, he's aware of all those things" | Patients, Family members, and Professionals |
| 28 | Word-finding | Cognitive functioning | Being able to find and say the correct word in context. | "Particularly with word finding, because that's so very frustrating for people" | Patients, Family members, and Professionals |
| 29 | Ability to assess risk | Cognitive functioning | Being able to spot potential dangers or hazards and make informed decisions by balancing whether someone has the right resources and abilities to complete a task without harming themselves or others. | "People lose sight of being able to risk assess" | Patients, Professionals |
| 30 | Adapting to change | Cognitive functioning | Being able to adjust to new situations, challenges, or conditions in a flexible and proactive way. | "Can't do change." | Patients, Professionals |
| 31 | Brain fog | Cognitive functioning | A non-medical term for a group of symptoms that affect your thinking, memory, and concentration. | "There is a brain fog that, I have had that." | Patients, Professionals |
| 32 | Semantic memory | Cognitive functioning | Being able to recall or recognise facts, meanings, and general knowledge. | "I find things that, that look vaguely familiar, and I think, what on Earth is that? What do you do with that? And I don't know what you do with it." | Patients, Professionals |
| 33 | Thinking or processing | Cognitive functioning | Being able to handle information and reflect on it. | "I'm a bit slower than I used to be, thinking wise" | Patients, Professionals |
| 34 | Effort and recovery time needed after completing a cognitively difficult task | Cognitive functioning | How tiring or effortful it is to attempt to complete a task (irrespective of success). | "I think there might actually be some mileage in that as a theme you know, to look at how much recovery time do people need after an activity." | Patients, Professionals |
| 35 | Ability to start a task (task initiation) | Cognitive functioning | Being able to start a project or task efficiently and on time without procrastination. | "When he's got going, you can't stop. But it's, it's the getting going." | Patients, Family members |
| 36 | Knowing or using strategies to help memory | Cognitive functioning | Being able to navigate memory difficulties using tools or methods (e.g., reminder systems). | "Like most people, you know, I have to put everything into my diary, you know, reminders. So, I've developed a whole range of “tactics”, if you like, to help me deal with the consequences of the condition." | Patients, Family members |
| 37 | Prospective memory | Cognitive functioning | Being able to "remember to remember". | "It's no good saying to him “Tomorrow can you unplug the… do something to the…?” you know? “Can you do such and such a thing tomorrow?” I have to think, “Oh, I need to ask him at the right time when I want him to do it”. Yeah. And so I need to make an aide-mémoire to myself, telling me to ask him at the right time." | Patients, Family members |
| 38 | Procedural memory | Cognitive functioning | Being able to implicitly remember how to do something, often referred to as "muscle memory". | "There's certain things that he has taught me how to do or told me, that he doesn't remember himself now." | Patients, Family members |
| 39 | Organisation | Cognitive functioning | Being able to arrange information, tasks, and items into a structure which is easy to manage. | "In fact, if you go and have a look now, it's a right mess. There's bits everywhere." | Patients, Family members |
| 40 | Perseverance or completing a task | Cognitive functioning | Continuing to something, despite facing difficulty or delays in achieving success. | "You get around every, you always have to get around things. You can't accept the status quo. So, I'll learn how to get around it, somehow or other. " | Patients, Family members |
| 41 | Planning | Cognitive functioning | Being able to think about the future, set a goal, and map out the steps to achieve it. | "I'll make a shopping list rather than just going to the supermarket and remembering what I need. I want, you know, now I put together a shopping list. You know, so there are. There are other examples of other situations where I now plan and prepare for things in a way that I never had to do before". | Patients, Family members |
| 42 | Spiritual wellbeing or connection to something bigger than you | Cognitive functioning | Having a sense of connectedness to something non-physical. For example, this could be a religion or God, to nature or the earth, or feeling there is meaning or purpose in life. | "What matters most to him is his spiritual life." | Patients, Family members |
| 43 | Motivation | Cognitive functioning | Having desire, enthusiasm, or drive to initiate and sustain goal-directed behaviours. | "I think “right, I’ll write down what I’ve got to do”. And then I'm trying to do everything I don't want to do and it's too hard." | Patients, Family members |
| 44 | Confidence in memory or cognition | Cognitive functioning | The degree of trust a person has in the accuracy or reliability of their own memory or cognition. | "It's a confidence thing, confidence that your memory is your memory. Yeah, and not having that. OK, I know this has happened or somebody's done this, but I'm not sure who it is." | Patients |
| 45 | Feeling confused | Cognitive functioning | Being unclear or uncertain on what is happening around you or what to do next. | "Just a bit of confusion at things." | Patients |
| 46 | Immediate (short-term) memory | Cognitive functioning | Being able to hold a small amount of information 'in mind' for a brief period of time. | "I had a feeling that I was perhaps, especially repeating myself... And [my wife] has just said my short-term memory wasn't very good." | Patients |
| 47 | Working memory | Cognitive functioning | Being able to hold and work with a small amount of information in mind in order to complete a task. | "I started to become aware that it was becoming more difficult because, as I say, it required you to have or required me to, to hold two or three different things in my mind." | Patients |
| 48 | Orientation | Cognitive functioning | Knowing where you are, what time/day/month it is, and who people around you are | "I never know what flipping day it was. It is. But then I suppose that's when you're not going to work or anything you can do." | Patients |
| 49 | Navigating | Cognitive functioning | Planning and following a route to get from one place to another. | "You know, geographically, driving a car or finding a way to places. I've, I think that's definitely got worse." | Patients |
| 50 | Associative memory or learning | Cognitive functioning | Learning and remembering links between pieces of information. | "It's that connection for me. I might remember half of something, but not the other half. But then not everybody's gonna be the same. So, it's things that connect, I suppose." | Patients |
| 51 | Spatial memory | Cognitive functioning | Being able to remember where things are in the environment and to plan a route to a location. | "He knew where places were in relationship to other places. Now, that's got a little bit more difficult." | Family members |
| 52 | Verbal fluency | Cognitive functioning | How quickly and easily a person can produce words, often from a given category. | "It doesn't have to be memory. It could be verbal fluency." | Professionals |
| 53 | Treatment adherence | Delivery of care | The extent to which participants adhered to the intervention protocol. | "Whether you are actually doing it, you know what I mean." | Patients, Professionals |
| 54 | Enjoyment of the intervention | Delivery of care | How enjoyable patients found the intervention itself. | "There is something to be gained in the fact that they enjoy it." | Professionals |
| 55 | Low mood | Emotional functioning/wellbeing | Feelings of being fed up, sad, or unhappy. Low mood is more short-term and less severe than depression. | "Perhaps something like mood" | Patients, Family members, and Professionals |
| 56 | Embarrassment due to cognitive difficulties | Emotional functioning/wellbeing | Feeling self-conscious or concerned that others may not perceive you well due to problems with your thinking or memory. | "I do think there's, some kind of, shame is probably a strong word, but some people feel embarrassed that they can't, their brain isn't as good as it was." | Patients, Family members, and Professionals |
| 57 | Frustration | Emotional functioning/wellbeing | Feelings of being upset or annoyed as a result of being unable to change or achieve something. | "It's just quite frustrating." | Patients, Family members, and Professionals |
| 58 | Self-confidence | Emotional functioning/wellbeing | Being able to trust in one's own abilities, qualities, and judgement. | "If there's any treatment that would give [my partner] back a bit more confidence". | Patients, Family members, and Professionals |
| 59 | Sense of agency, autonomy, or control | Emotional functioning/wellbeing | Feeling of being in control of one's actions and the external events that result from them. | "Control. Yeah, yeah. Of my life. Which is a pain." | Patients, Family members, and Professionals |
| 60 | Wellbeing or psychological health | Emotional functioning/wellbeing | Feeling “well” in yourself. For example, feeling good or positive about your life at the moment, and that you have a good quality of life. | "How's life? How's their wellbeing?" | Patients, Family members, and Professionals |
| 61 | Contentment | Emotional functioning/wellbeing | A state of happiness and satisfaction. | "Do I feel good? Do I feel content with life? Do I feel in decent spirits you know?" | Patients, Professionals |
| 62 | Hope | Emotional functioning/wellbeing | Expecting positive outcomes with respect to events and circumstances in one's life or the world. | "There's no hope. Yeah. Yeah. But if you feel like there's a bit of hope. Yeah. And I want to test that hope." | Patients, Professionals |
| 63 | Feeling able to cope | Emotional functioning/wellbeing | Being able to handle a challenging or stressful situation through aiming to resolve it or better handle the associated emotions. | "I'm just trying to cope with what I'm doing". | Patients, Family members |
| 64 | Rumination | Emotional functioning/wellbeing | Engaging in a repetitive negative thought process that loops continuously in the mind without end. | "Rumination is, is a good word for him." | Patients, Family members |
| 65 | Self-esteem | Emotional functioning/wellbeing | Feeling that you have value and personal worth. | "My daughter keeps saying to me “Mum, you're not stupid” because I keep saying, “oh, I'm stupid.” She says “no, you’re not. We all forget"." | Patients, Family members |
| 66 | Worry | Emotional functioning/wellbeing | Feeling troubled or preoccupied by events or outcomes that might happen in the future. | "It's just a bit of a worry, like, you know, thinking. You know. Could it get a lot, lot worse than what it is now?" | Patients, Professionals |
| 67 | Emotional lability | Emotional functioning/wellbeing | Experiencing rapid, uncontrolled, and exaggerated changes in moods. | "I didn't get upset so much with things like that. Because I get upset quite easily through no fault of anybody’s, it's just me." | Patients |
| 68 | Resilience | Emotional functioning/wellbeing | Being able to "bounce back" after challenging experiences, adapting and recovering from them. | "I think to keep going, that's about it. Just keep going. Just as they say, just keep paddling. Just keep paddling" | Patients |
| 69 | Stress | Emotional functioning/wellbeing | Negative state arising from perceiving or experiencing demands which exceed the skills or resources you have. | "It's about being able to live, you know, successfully independently. And minimising the, minimising any stress involved in doing those things" | Patients |
| 70 | Vulnerability | Emotional functioning/wellbeing | Being exposed to the risk of physical or emotional harm. | "I feel more fragile." | Patients |
| 71 | Adjustment to diagnosis | Perceived health status | Accepting and understanding what MCI is. | "I'm not sure she realises how much it affects me having to acknowledge that change." | Patients, Professionals |
| 72 | Staying in my own home | Personal circumstances | Living in the home, either supported or independently, rather than going into care. | "I think if the person feels that they're happily living at home and that they're safe." | Patients, Professionals |
| 73 | Balance or feeling steady | Physical functioning | Feeling that you can stand or sit without falling and maintaining your centre of gravity. | "I was wandering about across, you know, just unsteady. Maybe that's a word, the word describing it." | Patients, Professionals |
| 74 | Driving | Physical functioning | Retaining driving license and ability to drive. | "But I don't want to lose that bit of me that can drive. I do feel completely normal in it when I'm driving. It doesn't worry me, but if they do take that away from me, I should be quite upset, also because of where we live." | Patients, Family members, and Professionals |
| 75 | Gait | Physical functioning | How someone moves or walks. | "Certainly, I noticed during this year, my gait changed. But then I'm very conscious that there's a gait with Alzheimer's, you know, gait changes where you walk." | Patients, Family members, and Professionals |
| 76 | Maintaining independence (IADLs) | Physical functioning | Continuing to do activities which allow you to not rely on others such as paying your bills, cooking, going shopping, and using public transport. | "IADLs and ADLs. Yeah. So, whether the person can manage the things that they need to manage, whether they can perform all the aspects of life that they need to do, that's the critical thing." | Patients, Family members, and Professionals |
| 77 | Physical health or wellbeing | Physical functioning | The overall condition of the body, including how well your organs and systems function, and whether you have a health condition, injury, or illness. | "Keep healthy." | Patients, Professionals |
| 78 | Feeling tired or sleepy during the day | Physical functioning | Experiencing fatigue, sleepiness, or low energy during daytime hours. | "I would always have to stop because I was so tired. Close my eyes. Umm, not fall asleep, but I was just so exhausted. I just felt like my brain was tired" | Patients, Family members, and Professionals |
| 79 | Keeping physically active / exercising | Physical functioning | Being able to continue moving recreationally or for fitness. | "It's really important to keep active." | Patients, Family members |
| 80 | Managing personal care (basic ADLs) | Physical functioning | Continuing to do the day-to-day essentials of self-care such as eating, getting dressed, bathing or showering, and using the toilet. | "I think what’s more important to my patients is not how well they do on an esoteric cognitive test, more about how they are able to look after themselves and care for themselves." | Family members, Professionals |
| 81 | Falling | Physical functioning | Unintentionally ending up on the floor or a lower surface after losing balance. | "A sixty-seven-year-old chap shouldn't really be falling around the street" | Patients |
| 82 | Meeting social roles and expectations | Role functioning | Being able to participate in behaviours and fulfil responsibilities relating to their role (e.g., as a parent or partner). | "To do what I'm supposed to do." | Patients, Family members, and Professionals |
| 83 | Working | Role functioning | Being able to maintain employment. | "I wanted to go back to work really. And I just don't feel, and still don't feel, I can." | Patients, Family members, and Professionals |
| 84 | Ability to communicate | Social functioning | Being able to share information, ideas, or feelings with someone else so that you understand each other. Communication can be verbal (speaking), non-verbal (gestures, body language) or written. | "I do try to do that and if it's something that is important, if it counts, to spit it out, I'll find other ways to express myself." | Patients, Family members, and Professionals |
| 85 | Feeling stigmatised | Social functioning | Feeling devalued, rejected, or judged by others because of having MCI. | "People then sort of label you, don’t they?" | Patients |
| 86 | Regular social interactions | Social functioning | Spending time in the company of others and interacting with other people generally, as opposed to feeling socially isolated. | "So, I've sort of got drawn in on myself a bit. I’m quite a loner." | Patients, Family members, and Professionals |
| 87 | Meaningful relationships | Social functioning | Feeling that you have a good relationship and/or enjoy spending time with people you are close to and care about (e.g., with friends, family, a partner, or people in your community). | "My friendship with a whole load of other people, apart from my really very best mates, I find it a little bit challenged." | Patients, Family members, and Professionals |
| 88 | Participating in leisure or meaningful activities | Social functioning | Engaging in activities such as meeting friends, going to social clubs, or hobbies that are personally enjoyable or meaningful. | "I can't do what I want to do now." | Patients, Family members, and Professionals |
| 89 | Self-identity | Social functioning | Feeling like oneself and continuing to do activities that they enjoy and would normally do. | "You want to carry on being you, don't you?" | Patients, Family members, and Professionals |
| 90 | Tolerance of others | Social functioning | Being able to respect and accept the beliefs, feelings, habits, or behaviours of others, even if they differ from your own. | "His tolerance level of, of other people has changed." | Patients, Family members |
| 91 | Quality of life | Global quality of life | An assessment of overall wellbeing encompassing multiple domains including physical, mental, social, and environmental factors. | "I wouldn't say it's perfect, but it's good. You know the quality of my life is good enough, if you like. You know, it could be better, but it's good enough." | Patients |
| Resource use |  |  |  |  |  |
| 92 | Treatment costs | Economic | Monetary cost of intervention to the individual and/or organisation who provides it and whether the treatment provides reasonable value-for-money in terms of the benefits vs. costs. | "How much will it cost?" | Professionals |
| 93 | Effect on others/loved ones | Societal/carer burden | The amount of perceived or actual impact on others, including physical, emotional, social, and financial burden. | "Almost the last thing I want is to be in a situation where I would have to adapt how I live to fit in with somebody else. Because the routines and the things that the tactics I use, that would impinge on whoever I was living with." | Patients, Family members, and Professionals |
| Adverse events | |  |  |  |  |
| 94 | Side effects | Adverse events/effects | Unintended reaction or outcomes that occur in addition to the desired effect of the treatment or medication. | "I've spent my whole life trying to avoid medications because I've seen people having side effects." | Patients, Professionals |

#### Creating the ‘long list’ from outcomes identified during the review and qualitative interviews

Unique outcomes identified during the review and qualitative interviews were reviewed within the study management group

| **Review outcome(s)** | **Interview outcome(s)** | **Outcome included in the ‘long list’ for the Delphi survey** |
| --- | --- | --- |
| - | Survival | Survival |
| Adverse events | Side effects | Possible side effects |
| Immediate (short-term) memory | Memory (general); Immediate (short-term) memory | Short-term or immediate memory |
| Working memory | - | Working memory |
| Episodic memory; Memory; Associative memory; Logical memory; Everyday memory; Declarative memory; Traditional Chinese medicine symptoms of dementia | Episodic memory; Memory (general); Associative memory | Memory of experiences and events (episodic memory) |
| - | Procedural memory | Action-based memory (procedural memory) |
| Semantic memory; Associative memory; Declarative memory | Semantic memory; Spatial memory; Associative memory | Remembering facts (semantic memory) |
| Prospective memory; Everyday memory | Prospective memory | Remembering to remember (prospective memory) |
| Global cognition; Neuropsychology; Non-memory cognition; Everyday cognitive function; Traditional Chinese medicine symptoms of dementia | Cognition | Overall cognitive functioning |
| Executive function; Verbal fluency; Inhibitory control; Abstract thinking; Cognitive flexibility; Fluid intelligence; Goal-oriented rehabilitation | Executive function; Problem-solving; Ability to assess risk; Adapting to change; Ability to start a task (task initiation); Organisation; Perseverance or completing a task; Planning; Verbal fluency; | Executive functioning |
| Processing speed; Information processing | Thinking or processing; Brain fog; Feeling confused | Thinking clearly and quickly (information processing) |
| - | Navigating; Spatial memory | Being able to find your way around |
| Attention; Cognitive/perceptual load; Concentration | Attention | Attention |
| Visuospatial abilities; visuoconstructional abilities | Spatial memory; | Visual spatial abilities |
| Orientation; Traditional Chinese medicine symptoms of dementia | Orientation; Feeling confused | Being orientated to what's happening around you |
| Cognitive/perceptual load | Effort and recovery time needed after completing a cognitively difficult task | How much effort it is to do something requiring thinking or memory (cognitive load) |
| Word-finding; Verbal fluency | Word-finding; Verbal fluency | Word-finding ability |
| Language; Conversational ability | Language comprehension | Language comprehension |
| Language; Conversational ability | Language production | Language production |
| Apathy | Motivation | Feeling motivated |
| Subjective memory; Subjective cognitive functioning; Confidence in cognition and memory; meta-cognitive ability; perceived control over memory; Understanding of factors affecting memory | Subjective cognitive awareness; Confidence in memory or cognition; Embarrassment due to cognitive difficulties | How you think about your own cognitive functioning (metacognition) |
| Memory strategies; Understanding of factors affecting memory; Alcohol intake | Brain health; Knowing or using strategies to help memory; Presence of risk factors for dementia; Drinking in moderation; Having a healthy body weight or BMI | Knowing and doing what helps my brain health |
| Rumination | Rumination; Worry | Repetitive negative thoughts (rumination) |
| Adherence and tolerability; Satisfaction with the intervention; Satisfaction with outcomes; Acceptability and feasibility; Long-term maintenance of intervention; Willingness to continue with intervention | Treatment adherence; enjoyment of the intervention | Compliance and satisfaction with the intervention |
| Resilience to stress | Feeling able to cope | Ability to cope |
| Acceptance | Adjustment to diagnosis; Adapting to change | Adjustment to diagnosis |
| Resilience to stress | Resilience | Resilience |
| Stress | Stress | Stress |
| Helplessness | Vulnerability | Feeling vulnerable |
| - | Frustration | Feeling frustrated |
| Psychological wellbeing; Neuropsychology; Mood; Wellbeing; Apathy; Helplessness; Distress; Mindfulness | Wellbeing or psychological health; Contentment; Hope; Low mood; Worry; Embarrassment due to cognitive difficulties; Emotional lability; Tolerance of others | Overall emotional health or wellbeing |
| Participation in activities; Number of outdoor activities; Goal-oriented rehabilitation | Participating in leisure or meaningful activities | Participating in meaningful activities |
| - | Sense of agency, autonomy, or control | Sense of agency, autonomy, and/or control |
| - | Self-identity | Sense of self-identity |
| Self-confidence; Self-efficacy; Confidence in balance | Self-confidence; Confidence in memory or cognition; Feeling able to cope | Self-confidence |
| Self-esteem | Self-esteem; Embarrassment due to cognitive difficulties; | Self-esteem |
| Quality of life | Quality of life | Quality of life |
| Health-related quality of life | - | Health-related quality of life |
| Physical health; Body composition; Traditional Chinese medicine symptoms of dementia; Fibroblast growth factor-2; | Physical health or wellbeing; Having a healthy body weight or BMI; Presence of risk factors for dementia | Physical health |
| Psycho-motor skills; Visuoconstructional abilities; Reaction time | - | Psychomotor skills |
| Physical fitness; Physical activity levels; Physical functioning; Functional mobility; Number of outdoor activities; Muscular strength; Handgrip strength; Muscle endurance (lower limbs) | Keeping physically active / exercising | Mobility and physical fitness |
| Balance; Confidence in balance | Balance or feeling steady | Feeling steady (balance) |
| Falls | Falling | Falls |
| Walking; Gait | Gait | Gait (walking) |
| Basic activities of daily living; Functional ability; Severity of symptoms | Managing personal care (basic ADLs) | Managing personal care (basic activities of daily living) |
| Instrumental activities of daily living; Functional ability; Neuropsychology; Severity of symptoms | Maintaining independence (IADLs); Working; Staying in my own home | Maintaining independence (instrumental activities of daily living) |
| - | Driving | Ability to drive |
| Sleep quality; Traditional Chinese medicine symptoms of dementia | Sleep quality | Sleep |
| - | Feeling tired or sleepy during the day | Feeling tired or sleepy in the day |
| Occupational performance | Meeting social roles and expectations; Working | Meeting social roles and expectations of others |
| - | Feeling stigmatised | Feeling stigmatised |
| Conversational ability; Traditional Chinese medicine symptoms of dementia | Ability to communicate; Tolerance of others | Ability to communicate |
| Social participation | Regular social interactions | Having regular social contact |
| Satisfaction with relationships; Relationships with others | Having meaningful relationships; Tolerance of others | Satisfaction with relationships |
| Spiritual wellbeing; Mindfulness | Spiritual wellbeing or connection to something bigger than you | Spiritual wellbeing (or connection to something bigger than you) |
| Amyloid-beta burden; Tau burden; Cortisol; Brain-derived neurotrophic factor | Biomarkers of pathology/aetiology; Brain health | Fluid markers of brain health |
| Brain function; Functional connectivity; Resting state or spontaneous brain activity; Brain activation during a cognitive task; Brain metabolism; Brain perfusion and cerebrovascular health; EEG signatures | Brain activity; Brain health | Functional markers of brain health |
| Brain volume or structure | Brain volume; Brain health | Structural markers of brain health |
| Progression to dementia (incidence); Dementia severity; Severity of symptoms | Progression to dementia; | How many people developed dementia during the study (incidence) |
| Time to progression to dementia | Time to progression to dementia | Time to progression to dementia |
| Cytokines; C-reactive protein; Platelet factor 4; Peripheral blood monocytes; Phagocytic activity | Neuroinflammatory biomarkers | Markers of (neuro)inflammation |
| Cholesterol; Insulin; Insulin sensitivity; Haemoglobin A1c; Erythrocyte fatty acid membrane composition; Leptin; Adiposity; Vitamin B metabolism; Homocysteine; Omega-3 intake; Cortisol; Insulin-like growth factor 1; Alcohol intake | Nutritional biomarkers; Eating a healthy diet; Drinking in moderation | Markers of metabolic and nutritional health |
| Anxiety; Neuropsychiatric symptoms; Neuropsychology; Mood; Traditional Chinese medicine symptoms of dementia | Symptoms of anxiety; Worry | Symptoms of anxiety |
| Depression; Neuropsychiatric symptoms; Neuropsychology; Mood; Helplessness; Traditional Chinese medicine symptoms of dementia | Symptoms of depression; Low mood | Symptoms of depression |
| Behavioural and psychological symptoms of dementia (BPSD); Neuropsychiatric symptoms; Neuropsychology; Apathy; Behavioural disturbance; Traditional Chinese medicine symptoms of dementia | Emotional lability | Behavioural and psychological symptoms common in dementia |
| Cardiorespiratory fitness; Aerobic fitness; Cardiovascular health; Vascular endothelial growth factor; Blood pressure; Traditional Chinese medicine symptoms of dementia | - | Markers of cardiorespiratory health |
| Traditional Chinese medicine symptoms of dementia | Headaches | Headaches |
| Sensory functioning | Eyesight; hearing | Eyesight and hearing |
| Cortisol awakening response | - | Circadian rhythms |
| Telomere length | - | Telomere length |
| Caregiver burden | Effect on others/loved ones | Effects on people who support me |
| NA | Treatment costs | Treatment cost |

#### Outcomes included in the Delphi survey

Lay language was used throughout. Commonly used research or medical terminology were provided in parentheses where considered necessary for conveying meaning. Examples were based on discussions within the patient and public involvement group and real-life examples provided during the interviews. All outcomes were rated from 1 to 9 (1-3 = not essential, 4-6 = important but not essential, 7-9 = essential).

| Outcome included in the Delphi survey and lay definition | Examples given in the Delphi survey |
| --- | --- |
| 1. Overall cognitive functioning   How someone's brain works, as a whole, to help understand and interact with the world. | For example, this includes memory, attention, language, problem-solving, and thinking (altogether - rather than separately looking at these outcomes). |
| 2. Executive functioning   A set of cognitive skills that help someone to plan and manage tasks so that you can achieve goals, whether those are long-term goals or what they want to do in the next hour. | For example, executive functioning helps someone to make decisions, plan and organise things, adapt when things change or go wrong, and not act impulsively. |
| 3. Thinking clearly and quickly   Being able to come up with ideas, 'take things in', or respond to something without much difficulty. This is sometimes referred to as "information processing". It is the opposite of feeling like you have brain fog or can't think straight. | For example, this might look like being able to think through solutions to a problem or coming up with good ideas on what to do at the weekend. |
| 4. Attention   Being able to concentrate or focus on something without getting distracted. | For example, being able to follow along with a conversation and filter out noise in the background or being able to stay 'on task' without starting something else. |
| 5. Being orientated to what's happening around you/them   Knowing the answers to "where are you?", "who are you?", and "when is it?", at a given moment. | For example, knowing roughly what time of day it is (morning, afternoon, or evening), what the date is, and who the people around you are, and knowing where you are right now. |
| 6. Being able to find the way somewhere   Navigating surroundings in familiar and unfamiliar places so to get somewhere specific or achieve a goal. This might involve using landmarks, following directions, or read a map to do this. | For example, this might be finding the way to the toilet and back in an unfamiliar building, finding the right ward on a hospital, or taking a slightly different route because of a closed road. |
| 7. Visual-spatial ability   Being able to recognise or interpret things and how they relate to each other*.* | For example, visual-spatial skills are needed to visualise how parts fit together when putting together furniture or a puzzle, produce an accurate drawing, and solve jigsaw puzzles. |
| 8. Short-term or immediate memory   Being able to hold information in mind for a very short period of time (less than a minute). | For example, being able to repeat a short sentence back to someone or remembering a phone number long enough to dial it. |
| 9. Working memory   Like a 'mental workspace'. Ability to hold information in the mind and work with it to complete a task. This is similar to short-term memory, but includes doing something with the information being remembered. | For example, being able to follow the storyline of a television show or chapter of a book, remembering where you just put your keys down, and keeping track of a conversation so it's possible to join in. |
| 10. Memory of experiences and events (episodic memory)   Being able to remember specific events or experiences and details about them. This includes anything that happened more than a minute ago up to years or decades ago. | For example, being able to remember what happened yesterday, meeting someone for dinner last week and where this happened, details about the house where you grew up, or a list of words asked to remember a few minutes beforehand. |
| 11. Action-based memory (procedural memory)   Remembering how to perform tasks and actions learned before, without needing to consciously think about how to do it. This is sometimes known as "muscle memory". | For example, remembering how to do everyday tasks and habits such as making a cup of tea or coffee and doing household chores, and skills involved in playing musical instruments or sports. |
| 12. Remembering facts (semantic memory)   General knowledge, facts, and concepts which are not tied to a personal experience. Someone may not remember where they learned it, but they 'know' it. | For example, recalling historical facts, remembering someone's name, understanding what traffic signs mean, or knowing what a "bicycle" is. |
| 13. Remembering to remember (prospective memory)   Remembering to do something in the future that was planned. | For example, remembering appointments, to take medication on time, or to post a letter tomorrow. |
| 14. Language comprehension   Understanding what others have said or written down. | For example, being able to understand what someone is saying in a conversation and what words mean. |
| 15. Language production   Being able to come up with an idea and produce the words and phrases needed to share it, without many mistakes or pauses. | For example, this would include being able to say words, write them down, or sign them (if someone uses sign language). |
| 16. Word-finding ability   Being able to recall or come up with the right words in the right context. This is a more specific ability than language production. | For example, being able to say the word "dog", rather than describing it ("it's small, with four legs, barks") or saying a different word ("wolf", "cat"). |
| 17. Feeling motivated   Feeling that a drive, need, or want to do something or make a change. Motivation could come from enjoyment, feeling one 'has to' do it (obligation), or generally to achieve a personal goal. | For example, a goal of finishing off a project around the house, wanting to learn a new skill, or enjoying the feeling of helping others so wanting to volunteer. |
| 18. How much effort it takes to do something requiring thinking or memory (cognitive load)   How much mental effort is needed to complete a task. This is related but different to overall cognitive functioning, because it is about effort or burden, rather than performance or ability. | For example, how much effort it is to complete everyday tasks such as going to the shops or having a conversation. Someone may still be able to do these things, but how much does it require in terms of energy? |
| 19. Repetitive negative thoughts (rumination)   How often someone dwells or gets stuck thinking about something negative or distressing, often about themselves. | For example, repeatedly thinking about a bad argument had with a loved one or focusing for a long time and without coming up with solutions on being upset that something is difficult or no longer possible. |
| 20. How you think about and manage your cognitive functioning (meta-cognition)   Understanding about someone's own cognitive functioning (such as thinking, memory, and language) and what they can do to manage it better. | For example, this could be generally how good someone thinks their thinking and memory is, understanding that they have difficulty remembering things so they keep a calendar and diary, or that taking quick breaks during something difficult can help with concentration. |
| 21. Knowing and doing what helps brain health   Knowing what to do to keep the brain as healthy as possible and taking steps to do those things. | For example, finding out about high blood pressure and trying to get it under control, not smoking, eating a healthy diet, and doing things to stay active. |
| 22. Overall emotional health or wellbeing   Having a generally positive emotional state, feeling mentally well, and being able to manage emotions. | For example, this includes having awareness of feelings, finding healthy ways to manage negative emotions, and having a sense of meaning and purpose in life. |
| 23. Participating in meaningful activities   Continuing to be able to and doing activities which are personally enjoyable, meaningful, or otherwise positive and add something to life.  What these activities are will vary from person to person, but it is something that matters to them personally. | For example, this might include playing a favourite game or sport, reading books, going to a club, meeting up with friends for coffee, or spending time gardening or out in nature. |
| 24. Agency, autonomy, or control   Feeling that in control of life and being able to make choices about things that matter, without too much influence from others. | For example, this might mean deciding what each day will involve, being able to express needs in a relationship, or being able to make decisions that affect healthcare. |
| 25. Self-identity   How you think about yourself in terms of who you are. | For example, this might include having preferences and values, personality, and the roles some has in a community or relationships. |
| 26. Self-confidence   Trust in oneself to handle different situations and challenges. This includes belief in abilities, qualities, strengths, skills, and judgement. | For example, being confident to participate in social activities, taking on a new project at home, or finding the way to a new cafe to meet a friend. |
| 27. Self-esteem   How you feel about yourself and your worth, regardless of abilities or achievement. | For example, this includes accepting and valuing yourself as you are instead of feeling bad about yourself, and not needing lots of reassurance from others about your self-worth. |
| 28. Ability to cope   Feeling able to cope and having strategies and behaviours to manage stress, challenges, and difficult emotions. Coping mechanisms can be positive or negative. | For example, positive coping strategies include exercise and seeking support, whilst negative coping strategies include isolation or substance use (alcohol or drugs). |
| 29. Adjustment to diagnosis   Accepting and adjusting to having a diagnosis of mild cognitive impairment and the symptoms that are part of it. | For example, this might include managing initial shock or disbelief, seeking information, taking steps to improve brain health, or coming up with strategies that help manage symptoms. |
| 30. Resilience   Being able to adapt and recover, grow, or 'bounce back' after challenging situations or stress. | For example, this might include being open to change, being optimistic, managing emotions, problem-solving, and continuing on with life after receiving bad news. |
| 31. Stress   The body's response to feeling threatened or pressured. Some stress can help you deal with challenges, whilst too much stress can be harmful. | For example, someone might experience stress because of financial worries, life changes such as moving house or losing a loved one, or from managing multiple health conditions needing lots of hospital appointments. |
| 32. Feeling vulnerable   Feeling exposed or sensitive to harm or being hurt in some way. | For example, people can feel emotionally or socially vulnerable (fear of rejection or judgement) or physically vulnerable (feeling physically unsafe or at risk of harm). |
| 33. Feeling frustrated   Feeling annoyed, upset, or discouraged because you cannot do something or because things haven't gone as planned. | For example, feelings of frustration might come from frequently losing items around the house, not being able to find the right word, or finding tasks that used to be easy more difficult. |
| 34. Symptoms of anxiety   Anxiety or worry is usually a natural response to feeling pressured, afraid, or threatened. It can show up in how we feel physically, mentally, and in how we behave. It can be mild or severe. | For example, feeling or worrying that something bad is going to happen, feeling restless, nervous, tense, or irritable, sweating, having a fast heartbeat, or having troubling relaxing. |
| 35. Symptoms of depression   Signs which might indicate low mood or depression. These involve changes in mood, thoughts, and behaviours and can vary in intensity and duration. | For example, feeling sad, irritable or empty, feeling hopeless or tearful, having too much or too little food, and feeling unable to concentrate. |
| 36. Behavioural and psychiatric symptoms associated with dementia (BPSD)   Behavioural, psychological, and emotional symptoms which are often seen in dementia. Please note that not everyone who has mild cognitive impairment will develop dementia. | For example, this includes anxiety and depression, but also hallucinations (seeing things that aren't there) and delusions (believing things that aren't true), agitation, wandering, and acting impulsively. |
| 37. Meeting social roles and expectations of others (role functioning)   Feeling able to participate in behaviours and fulfil responsibilities relating to their 'role' or in a specific relationship. | For example, feeling able to be a good partner, friend, employee, or parent and do what is expected in these roles. |
| 38. Ability to communicate   Being able to share information, ideas, or feelings with someone else to understand each other. Communication can be verbal (speaking), non-verbal (gestures, body language) or written. | For example, being able to clearly express thoughts and feelings, keeping on topic in conversation, or reacting appropriately to what someone else does or says. |
| 39. Having regular social contact   Spending time in the company of others and interacting with other people generally. This is the opposite of being socially isolated. | For example, this might include frequently texting or talking to others on the phone, chatting with people when out, or spending time in community settings. |
| 40. **Satisfaction with relationships** Having a good relationship and/or enjoy spending time with people who matter and you care about. | For example, this could be having good friends that you see often enough, getting along well with family, having a strong relationship with a partner where you feel happy and supported, or being part of a community group and feeling included and valued. |
| 41. **Feeling stigmatised** Feeling that you are treated unfairly or differently because of having mild cognitive impairment. | For example, someone might feel they are being left out of important decisions or that they are viewed as careless or mean when they forget something they've been told before. |
| 42. **Spiritual wellbeing (or connection to something beyond yourself)** Feeling a sense of peace, purpose, or connection to something bigger than oneself. Spiritual wellbeing involves having values, beliefs, and/or practices which help to provide meaning or direction in life. | For example, feeling connected to nature, mindfulness or inner peace, having a sense of purpose, engaging in religious practice or communicating with God or gods. |
| 43. **Physical health** The general 'condition' of someone's body - whether someone is well (or not unwell). | For example, this includes how well tissues, organs, and cells are functioning, as well as not having other health conditions, illnesses or injuries (these are sometimes called comorbidities). |
| 44. **Mobility and physical fitness** Being able to move freely and being physically strong enough to do everyday things. | For example, being able to reach for things, walk up and down stairs, carry heavy shopping, or do a favourite form of exercise without pain or discomfort. |
| 45. **Psycho-motor skills** Being able to coordinate mental processes with physical actions. This is most often done using hand-eye coordination but can involve other parts of the body. | For example, being able to grasp objects, typing on a keyboard, riding a bike, tapping a finger to a rhythm, or using a pool cue to hit a ball. |
| 46. **Gait**  How someone moves or walks. | For example, rhythm and speed of walking and whether they are balanced when moving. |
| 47. **Feeling steady (balance)** Being able to keep the centre of gravity (or stay upright) whilst moving or standing still. | For example, this includes keeping balanced whilst walking and recovering after a stumble to prevent falling, and not feeling dizzy or off-balance when standing. |
| 48. **Falls** How much someone falls over (that is, unintentionally and suddenly go down to the ground, usually from standing). | For example, how much someone trips over things or trips over when there was nothing in the way. |
| 49. **Managing personal care (basic activities of daily living)** Continuing to do the day-to-day essentials without support. | For example eating, getting dressed, bathing or showering, and using the toilet. |
| 50. **Maintaining independence (instrumental activities of daily living)** Continuing to do activities which allow someone to not rely on others and/or enable living independently. | For example, paying bills, cooking, going shopping, and using public transport. |
| 51. **Ability to drive** Being able to continue driving safely. | For example, some people can no longer drive because their mild cognitive impairment affects their concentration or reaction time to a hazard in the road. |
| 52. **Sleep** Quality and quantity of sleep. | For example, if you are having enough sleep, if your sleep is broken by lots of waking during the night, or if you don't have enough deep sleep. |
| 53. **Feeling tired and sleepy during the day** Feeling that someone could, or wants to, fall asleep during the daytime or feeling low in energy. | For example, this includes accidentally dozing off when sat down, feeling too tired to concentrate, or needing to go for a nap in the middle of the day. |
| 54. **Structural biomarkers of brain health** Outcomes relating to the size and shape of the brain as a whole and different parts of it. These are often measured using imaging techniques such as MRI (magnetic resonance imaging). | For example, looking at the size of the brain or specific regions in the brain or looking at connections between brain areas. |
| 55. **Functional biomarkers of brain health** Outcomes relating to brain function or activity. These are often assessed using neuroimaging techniques such as functional MRI or electrophysiology such as EEG (electroencephalography). | For example, measuring brain activity at rest or whether areas of the brain thought to be involved in memory are more or less active doing a memory task. |
| 56. **Fluid biomarkers of brain health** Indicators of brain health or disease found in bodily fluids such as the blood or cerebrospinal fluid (CSF). | For example, this might include proteins which are markers of Alzheimer's disease such as amyloid-beta or tau pathology, or markers of damage to brain cells such as neurofilament light chain (NFL). |
| 57. **How many people developed dementia during the study** The number of people with mild cognitive impairment who progressed to having dementia during the course of the study. *Please note that not everyone who has mild cognitive impairment will develop dementia.* | For example, 1 in 10 people who had treatment A developed dementia, compared to 1 in 100 people who had treatment B. |
| 58. **Time to progression to dementia** Whether having a treatment was linked to being diagnosed with dementia later in the study. *Please note that not everyone who has mild cognitive impairment will develop dementia.* | For example, people on treatment A on average developed dementia after having mild cognitive impairment for 5 years whilst people on treatment B on average developed dementia after 10 years. |
| 59. **Survival** Continuing to be alive. | For example, how a study affects life expectancy or whether more people on treatment A compared to treatment B are alive after 10 years. |
| 60. **Possible side effects** Unintentional effects of a treatment, that are often negative. | For example, a therapy intended to improve cognition might make someone more anxious and aware of their difficulties, or a drug to help thinking and memory might make someone feel sick or have a headache. |
| 61. **Markers of inflammation** Outcomes which suggest an immune or inflammatory response. Cells which are damaged, infected, or dying release signals which can be measured, often in blood. | For example, proteins such as C-reactive protein and cytokines are released in response to infection, inflammation, or disease in the body, including the brain and nervous system. |
| 62. **Markers of metabolic health and nutrition** Metabolism is how the body's cells are able to change food into energy. Nutrition is about getting the right nutrients (vitamins and minerals, water, carbohydrates, proteins, fats) to be healthy. | For example, markers of metabolic and nutritional health include blood sugar, cholesterol, and waist size. |
| 63. **Markers of cardiorespiratory health** Outcomes relating to the health of the heart and lungs. | For example, this could be heart rate, blood pressure, or how much oxygen is used during intense exercise (VO2 max). |
| 64. **Headaches** A pain in the head or face, which can last minutes, hours, or days and can be mild or severe. There are different types of headaches. | For example, some people feel pressure across the forehead (tension), others have throbbing on one side often above or behind one eye (migraine) which can make them feel sick.  Some people get them after doing certain activities. |
| 65. **Vision and hearing** Vision is being able to see and perceive colours, shapes, and patterns. Hearing is about perceiving sounds. Sensory information comes from the eyes and ears and is interpreted by the brain. | For example, how well someone can see at different distances and how well someone can hear sounds at different frequencies (tones). |
| 66. **Telomere length** The length of protective caps at the end of the chromosomes. They are made of DNA sequences which shorten each time a cell divides. Telomere length is an established marker of biological ageing (how well a body is functioning). | For example, someone may be 60 years old but have the telomere length of a 50 year old because they exercise, eat a healthy diet, don't smoke, get enough sleep, and manage their stress. Someone who is 60 but doesn't do any of these protective things might have shorter telomeres, similar in length to a 70 year old. |
| 67. **Circadian rhythm** The natural 24-hour cycle of physical, mental, and behavioural changes that happen in your body. | For example, this might be checking if your levels of melatonin increase throughout the evening to help you feel sleepy towards bedtime, or that your cortisol increases shortly after you get out of bed to help you feel awake. As well as sleep and waking, circadian rhythms affect eating behaviours, temperature, and blood pressure. |
| 68. **Quality of life** How good an individual's life is overall. Anything that can alter satisfaction with life is included. | For example, quality of life might be affected by health, relationships, and finances. |
| 69. **Health-related quality of life** The impact of health on quality of life. | For example, this includes how physical and mental health conditions and any treatments affect how you feel about life and how good it is. |
| 70. **Effects on people who support the person living with mild cognitive impairment** How much someone feels mild cognitive impairment has an effect on people who support them. | For example, feeling that cognitive impairment means others get annoyed with them, have to do more, or might worry others if they knew about it or noticed it. |
| 71. **Treatment cost** How much the treatment costs. | For example, this might be the cost per person per year of treatment to the NHS for a drug, or cost to the patient for taking part in a group therapy. |
| 72. **Satisfaction and engagement with treatment** Outcomes relating to how people perceived the treatment itself and whether people followed the treatment as it was supposed to be done. | In research, this is sometimes referred to as feasibility, acceptability, or compliance. |

#### Outcome scoring during the Delphi study

|  | Delphi Round 1 | | | Delphi Round 2 | | |
| --- | --- | --- | --- | --- | --- | --- |
|  | Patients | Family members | Professionals | Patients | Family members | Professionals |
| Overall cognitive functioning* | 8 | 8 | 9 | 8 | 8 | 9 |
| Executive functioning* | 8 | 8 | 8 | 8 | 8 | 8 |
| Thinking clearly and quickly | 7 | 7 | 7 | 7 | 7 | 6.5 |
| Attention* | 8 | 7 | 7 | 8 | 7 | 7 |
| Being orientated to what's happening around you/them (orientation)* | 8 | 8 | 8 | 8 | 8 | 8 |
| Being able to find the way somewhere (navigation)* | 7 | 6 | 7 | 7 | 6 | 7 |
| Visual-spatial ability | 6 | 5 | 7 | 7 | 5 | 6 |
| Short-term or immediate memory* | 8 | 7 | 8 | 8 | 7 | 7 |
| Working memory* | 8 | 7 | 8 | 8 | 7 | 7 |
| Memory of experiences and events (episodic memory)† | 7 | 6 | 7 | 7.5 | 5 | 7 |
| Action-based memory (procedural memory)* | 7 | 7 | 8 | 8 | 7 | 7 |
| Remembering facts (semantic memory) | 7 | 5 | 6 | 7 | 5 | 6 |
| Remembering to remember (prospective memory) * | 8 | 7 | 7 | 7 | 7 | 7 |
| Language comprehension* | 8 | 7 | 8 | 7 | 7 | 7 |
| Language production | 8 | 6 | 8 | 8 | 6 | 7 |
| Word-finding ability | 7 | 5 | 7 | 7 | 5 | 7 |
| Feeling motivated | 7 | 5 | 6 | 7 | 5 | 6 |
| How much effort it takes to do something requiring thinking or memory (cognitive load) | 8 | 6 | 6 | 7 | 6 | 5.5 |
| Repetitive negative thoughts (rumination) | 7 | 6 | 5 | 7 | 6 | 5 |
| How you think about and manage your cognitive functioning (meta-cognition) | 7 | 6 | 6 | 7 | 6 | 5.5 |
| Knowing and doing what helps brain health | 8 | 7 | 6 | 8 | 7 | 6 |
| Overall emotional health or wellbeing* | 8 | 7 | 7 | 8 | 7 | 7 |
| Participating in meaningful activities* | 9 | 7 | 7 | 8 | 7 | 7 |
| Agency, autonomy, or control | 8 | 7 | 7 | 8 | 7 | 7 |
| Self-identity | 8 | 6 | 6 | 7 | 6 | 5.5 |
| Self-confidence | 8 | 7 | 6 | 8 | 7 | 6 |
| Self-esteem | 8 | 6 | 5 | 7 | 6 | 5 |
| Ability to cope | 8 | 7 | 6 | 8 | 6 | 5 |
| Adjustment to diagnosis | 9 | 6 | 5 | 8 | 6 | 6 |
| Resilience | 8 | 6 | 5 | 7 | 6 | 5 |
| Stress | 8 | 6 | 5 | 7 | 5 | 5 |
| Feeling vulnerable | 7 | 6 | 5 | 7 | 6 | 5 |
| Feeling frustrated | 7 | 6 | 5 | 7 | 6 | 5 |
| Symptoms of anxiety* | 7 | 7 | 7 | 7 | 7 | 7 |
| Symptoms of depression* | 8 | 7 | 7 | 7 | 7 | 7 |
| Behavioural and psychiatric symptoms associated with dementia (BPSD) | 8 | 7 | 7 | 7 | 7 | 7 |
| Meeting social roles and expectations of others (role functioning) | 7 | 6 | 6 | 8 | 5 | 6 |
| Ability to communicate* | 8 | 8 | 7 | 8 | 8 | 7 |
| Having regular social contact | 8 | 7 | 6 | 8 | 7 | 6 |
| Satisfaction with relationships | 8 | 6 | 6 | 8 | 7 | 6 |
| Feeling stigmatised | 6.5 | 6 | 6 | 6 | 6 | 5 |
| Spiritual wellbeing (or connection to something beyond yourself) | 6 | 5 | 4 | 5 | 4 | 4 |
| Physical health* | 8 | 7.5 | 7 | 8 | 8 | 7 |
| Mobility and physical fitness | 8 | 6 | 7 | 8 | 6 | 6 |
| Psycho-motor skills | 7 | 7 | 6 | 7 | 7 | 6 |
| Gait | 6.5 | 6 | 6 | 6 | 6 | 6 |
| Feeling steady (balance) | 7.5 | 7 | 6 | 7 | 7 | 6 |
| Falls | 7 | 7 | 6 | 7 | 7 | 6 |
| Managing personal care (basic activities of daily living)* | 8 | 8 | 8 | 8 | 8 | 8 |
| Maintaining independence (instrumental activities of daily living)* | 8 | 7 | 9 | 8 | 7 | 9 |
| Ability to drive | 7.5 | 6 | 6 | 7 | 5 | 6 |
| Sleep* | 8 | 7 | 7 | 8 | 7 | 7 |
| Feeling tired and sleepy during the day | 7 | 6 | 6 | 7 | 6 | 6 |
| Structural biomarkers of brain health* | 8 | 7 | 7 | 8 | 7 | 7 |
| Functional biomarkers of brain health | 8 | 7 | 6 | 8 | 7 | 6 |
| Fluid biomarkers of brain health* | 9 | 8 | 6 | 8 | 8 | 7 |
| How many people developed dementia during the study* | 9 | 7 | 8 | 8 | 7 | 8 |
| Time to progression to dementia* | 9 | 8 | 8 | 9 | 8 | 8 |
| Survival | 8 | 7 | 7 | 8 | 7 | 8 |
| Possible side effects‡ | 7 | 6 | 8 | 7 | 6 | 8 |
| Markers of inflammation | 8 | 7 | 6 | 8 | 7 | 6 |
| Markers of metabolic health and nutrition | 7 | 6 | 6 | 7 | 6 | 6 |
| Markers of cardiorespiratory health | 8 | 6 | 6 | 7 | 6 | 5 |
| Headaches | 7 | 6 | 5 | 7 | 6 | 5 |
| Vision and hearing | 8 | 7 | 6 | 8 | 6 | 5 |
| Telomere length | 8 | 6 | 4 | 7 | 5 | 4 |
| Circadian rhythm | 8 | 6 | 5 | 7 | 5 | 5 |
| Quality of life* | 8 | 8 | 9 | 8 | 8 | 8 |
| Health-related quality of life* | 8 | 7 | 8 | 7 | 8 | 8 |
| Effects on people who support the person living with mild cognitive impairment* | 8 | 7 | 8 | 7 | 7 | 8 |
| Treatment cost | 7 | 5 | 6 | 6 | 5 | 6 |
| Satisfaction and engagement with treatment‡ | 8 | 6 | 7 | 7 | 6 | 7 |

During the Delphi study, each of the below 72 outcomes were rated by participants. Median scores for each outcome are provided for each stakeholder group for round one and two. **Key:** * = included within final core outcome set; † = added during consensus meeting; ‡ = re-evaluated during consensus meeting as not relevant to effectiveness.

#### Outcome merging through Delphi study to Consensus Meeting

Table of outcomes included by consensus in round two of the Delphi survey and how outcomes were included in the final core outcome set. The first column lists the lay outcome name used in the Delphi survey. Due to a high number of outcomes included at this stage, the study management group met to merge outcomes based on conceptual similarity and pragmatism (i.e., whether typically outcome measurement tools would measure these concepts together). Proposed merged outcomes were reviewed and discussed at the consensus meeting with stakeholders. Where changes were made during the consensus meeting, this is emphasised in **bold** text.

| DELPHI SURVEY OUTCOME NAME | PROPOSED MERGED OUTCOME | REVISED FINAL OUTCOME | NOTES |
| --- | --- | --- | --- |
| Executive functioning | Cognitive functioning | COGNITIVE FUNCTIONING (NON-MEMORY) | Consensus meeting participants recognised the importance of cognitive function in MCI. Given the breadth of cognitive functions identified as essential in the Delphi study, a single cognitive functioning outcome was considered insufficient.  **Cognitive functioning was split into memory and non-memory** cognitive functions in recognition of the recognised amnestic and non-amnestic symptoms and sub-types of MCI.  The domains here should all be included within one or more cognitive assessments. |
| Attention |  |  |  |
| Being orientated to what’s happening around you/them (Orientation) |  |  |  |
| Being able to find the way somewhere (Navigation) |  |  |  |
| Language comprehension |  |  |  |
| Short-term or immediate memory |  | MEMORY |  |
| Working memory |  |  |  |
| Action-based memory  (Procedural memory) |  |  |  |
| Remembering to remember (Prospective memory) |  |  |  |
| Overall emotional health and wellbeing | Emotional functioning/wellbeing | MENTAL HEALTH AND WELLBEING | There was discussion about terminology to use that encompasses both positive and negative aspects of mental and emotional health, but there was overall agreement that these concepts overlapped considerably and could be considered one outcome. |
| Symptoms of anxiety | Psychiatric outcomes |  |  |
| Symptoms of depression |  |  |  |
| Effects on people who support the person living with MCI | Social functioning/relationships | SOCIAL FUNCTIONING/RELATIONSHIPS | No change, though multiple other similar outcomes were put forwards for voting (e.g., having regular social contact, role functioning). |
| Ability to communicate |  |  |  |
| Structural biomarkers of brain health | Biomarkers of brain health | BIOMARKERS OF BRAIN HEALTH | No change. The panel were cognisant that measuring biomarkers is complicated by practical constraints (e.g., funding, access) and changes to best practice over time (e.g., emergence of new biomarkers) and recommended pragmatism. |
| Fluid biomarkers of brain health |  |  |  |
| How many people developed dementia during the study | (Likelihood of) progression to dementia | PROGRESSION TO DEMENTIA | **Participants felt removing ‘(Likelihood of)’ had greater clarity** and agreed that incidence and time to progression could be collapsed into a single merged outcome. |
| Time to progression to dementia |  |  |  |
| Managing personal care (ADLs) | Independence/functional ability | EVERYDAY FUNCTIONING AND INDEPENDENCE | Participants agreed that these outcomes reflected different levels of physical functioning/activities of daily living but **preferred the term ‘everyday functioning and independence’** as ‘functional ability’ was considered ambiguous. |
| Maintaining independence (IADLs) |  |  |  |
| Participating in meaningful activities |  |  |  |
| Physical health | General/physical health | GENERAL/PHYSICAL HEALTH | No change. |
| Sleep | Sleep | SLEEP | No change. The panel was asked whether this could be meaningfully collapsed within everyday general/physical health, but this was not considered specific enough. |
| Quality of life | Global quality of life | QUALITY OF LIFE | Participants felt that health-related quality of life was a subcategory of global quality of life and so should be considered one outcome. |
| Health-related quality of life | Health-related quality of life |  |  |
| Satisfaction and engagement with the treatment | - | | Keep outcome name as-is but was removed from the COS as it was not considered an effectiveness outcome nor of specific relevance to MCI. |
| Possible side effects | - | | Keep outcome name as-is but was removed from the COS as it was not considered an effectiveness outcome nor of specific relevance to MCI. |

The following outcomes were ‘undecided’ during the Delphi study. The consensus panel was asked to nominate outcomes to be put forward for voting for inclusion in the final COS. Only one outcome, *Memory of experiences or events (episodic memory*), reached consensus to be included in the final COS.

Two participants from the panel (one patient, one family member) were unavailable during the end of the voting period due to personal circumstances but supported the final COS via email.

| DELPHI SURVEY OUTCOME NAME | FINAL DECISION | VOTING AGREEMENT |
| --- | --- | --- |
| Memory of experiences or events (episodic memory) | IN – included as part of ‘Memory’ outcome. | 13/13 (100%) |
| Remembering facts (semantic memory) | OUT |  |
| Visual-spatial ability | OUT |  |
| How much effort it takes to do something requiring thinking or memory (cognitive load) | OUT | 1/13 (8%) |
| Thinking clearly and quickly | OUT |  |
| Language production | OUT |  |
| Word-finding ability | OUT |  |
| Feeling motivated | OUT |  |
| How you think about and manage your cognitive functioning (meta-cognition) | OUT |  |
| Knowing and doing what helps brain health | OUT |  |
| Feeling frustrated | OUT | 2/13 (15%) |
| Ability to cope | OUT |  |
| Adjustment to diagnosis | OUT | 5/13 (38%) |
| Resilience | OUT | 6/13 (46%) |
| Stress | OUT | 1/13 (8%) |
| Feeling vulnerable | OUT |  |
| Self-identity | OUT |  |
| Self-confidence | OUT |  |
| Self-esteem | OUT |  |
| Agency, autonomy, or control | OUT |  |
| Repetitive negative thoughts (rumination) | OUT |  |
| Meeting social roles and expectations of others (role functioning) | OUT | 5/11 (45%) |
| Having regular social contact | OUT | 6/11 (55%) |
| Satisfaction with relationships | OUT |  |
| Feeling stigmatised | OUT |  |
| Spiritual wellbeing (or connection to something beyond yourself) | OUT |  |
| Functional biomarkers of brain health | OUT |  |
| Behavioural and psychiatric symptoms associated with dementia | OUT | 2/11 (18%) |
| Ability to drive | OUT |  |
| Mobility and physical fitness | OUT |  |
| Psycho-motor skills | OUT |  |
| Markers of inflammation | OUT |  |
| Markers of metabolic health and nutrition | OUT |  |
| Markers of cardiorespiratory health | OUT |  |
| Telomere length | OUT |  |
| Headaches | OUT |  |
| Vision and hearing | OUT |  |
| Gait | OUT |  |
| Feeling steady (balance) | OUT |  |
| Falls | OUT |  |
| Feeling tired and sleepy during the day | OUT |  |
| Circadian rhythm | OUT | 1/11 (9%) |
| Survival | OUT |  |
| Treatment cost | OUT |  |
